# Direct and mediated effects (DME) SLCMA: a novel method for life course modelling with time-varying covariates

**DOI:** 10.64898/2026.05.29.26354427

**Authors:** Solomon Beer, Andrew J. Simpkin, Nicola Fitz-Simon, Sherief Y. Eldeeb, Heather J Zar, Dan J Stein, Erin C. Dunn, Andrew D.A.C. Smith

**Affiliations:** School of Mathematical and Statistical Sciences, University of Galway, Galway, Ireland; Department of Sociology, College of Liberal Arts, Purdue University, West Lafayette, IN; Center for Genomic Medicine, Massachusetts General Hospital, Boston, MA; SAMRC Unit on Child and Adolescent Health, Department of Paediatrics and Child Health, Faculty of Health Sciences, University of Cape Town, Cape Town, South Africa; SAMRC Unit on Risk and Resilience in Mental Disorders, Department of Psychiatry and Neuroscience Institute, University of Cape Town, Cape Town, South Africa; Department of Psychiatry, Harvard Medical School, Boston, MA; Mathematics and Statistics Research Group, University of the West of England, Bristol, UK

**Author notes:** Correspondence: Address correspondence to: Andrew D.A.C. Smith, PhD, MStats, BSc, Associate Professor of Statistics, FET - Computer Science and Creative Technologies, University of the West of England, Bristol, United Kingdom, BS16 1QY.

**Keywords:** Life course modelling, structured approach, time-varying covariates, DCHS

## Abstract

**Background:** In prospective cohort studies, where an exposure is collected repeatedly, interest often lies in determining whether the timing of that exposure has a differential effect on a later outcome. The Structured Life Course Modeling Approach (SLCMA), where users select between temporal hypotheses of exposure specified a priori, provides one way to analyse such longitudinal data. However, few studies using SLCMA consider the effect of time-varying covariates (TVC) which may impact associations.

**Methods:** We present a modified version of the SLCMA – called direct and mediated effects (DME)-SLCMA – which corrects for TVC. We first develop the DME-SLCMA method, test it through simulation, and apply it to psychosocial data from the Drakenstein Child Health Study (DCHS, *n*=336) to investigate relationships between maternal psychopathology, TVC of socioeconomic status, and offspring depressive symptoms.

**Results:** We found that, on average, offspring depressive symptoms score increased by 3.9% (95% CI: 1.0%-6.9%, *p* = 0.039) for each unit of maternal psychopathology (SRQ) at 48 months whilst adjusting for time-varying socioeconomic status (at 18, 30, 42 and 54 months). Our simulations identified several realistic scenarios where selections ignoring TVC - with TVC mediated exposure effects present - were prone to be incorrect, including our DCHS example.

**Conclusion:** DME-SLCMA is a robust new approach for life course modelling in the presence of time-varying covariates. We recommend adjusting for TVC whenever possible, and, when not possible, our simulation study identified that scenarios where mediated effects are comparable, or greater, in magnitude to direct effects are most prone to confounding.

## Introduction

One strength of longitudinal studies is the availability of repeated measurements of an exposure prior to a later-life outcome, enabling investigation across the life course rather than a narrow focus on age-specific effects. Such repeated measures allow the investigation of structured hypotheses regarding this life course relationship^1,2^.

The structured life course modelling approach (SLCMA)^3–5^ has been established as a method for identifying and testing these hypothesized relationships. Smith et al. (2016) ^6^ proposed methods for adjusting for confounders (i.e. covariates measured at baseline) within the SLCMA. However, the same longitudinal studies that have repeated measurements of exposures are likely to have repeated measurements of covariates. Yet, no studies to our knowledge have applied the SLCMA with time-varying covariates. In this study, we propose a novel approach for such adjustment.

We examined the effect of maternal psychopathology on offspring using data collected when they were 12-48 months on depressive symptoms at 60 months of age. Data came from the Drakenstein Child Health Study (DCHS)^7,8^ a longitudinal cohort study sampling predominantly Black and Coloured population groups in South Africa (see supplementary materials for commentary on ethnicity category usage). Maternal psychopathology was measured using the Self-Reporting Questionnaire-20 (SRQ-20) ^9^ and provides a score which comprises of depression and anxiety components, with a mean prevalence across time-points of 2.51 (2.96). In DCHS, socio-economic status (SES) was also repeatedly measured at 18, 30, 42 and 54 months. The depressive symptoms score used, was the natural log of the externalising problems subscore of the Child Behavioural CheckList (CBCL)^10,11^ measured at 60 months. Further details on these variables are available in the **Supplementary Information**.

**Figure 1** shows a potential causal directed acyclic graph (DAG) with repeated measurements of both exposure (maternal psychopathology) and covariate (SES).

**Figure 1.**
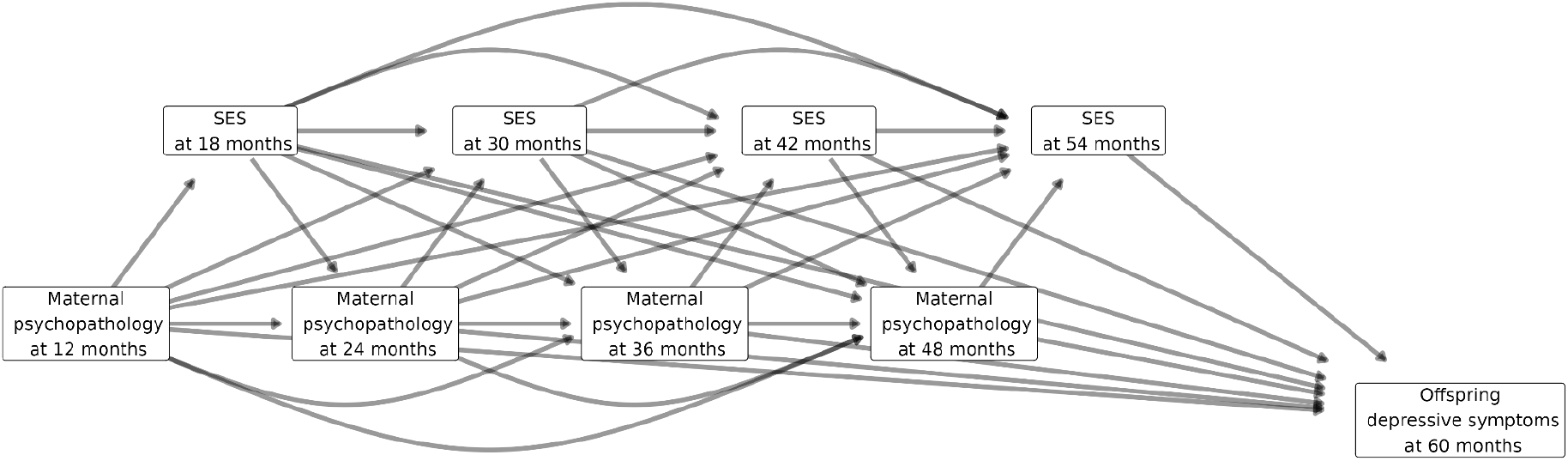
Directed acyclic graph example with a repeatedly-measured exposure (maternal psychopathology) and a repeatedly-measured time-varying covariate (socioeconomic status) and their effect upon offspring depressive symptoms.

**Figure 1** depicts two ways a time-varying covariate can relate to the exposure-outcome relationship across the life course. Firstly, a covariate can act as a confounder by having a causal effect on the outcome, and the exposure at a later timepoint; in this scenario statistical adjustment for the covariate helps reduce confounding bias. Secondly, a covariate can act as a mediator by having a causal effect on the outcome, and being causally affected by the exposure at a preceding timepoint; in this scenario adjusting for the covariate discounts the indirect or mediated effect of the exposure. Careful thought is therefore required regarding whether, and how, to adjust for a time-varying covariate in the SLCMA. In this paper we propose the direct and mediated effects (DME) SLCMA – pronounced “dee-mee-slick-mah” – as a suitable approach for investigating structured life course hypotheses in the presence of time-varying covariates.

### DME SLCMA

Our model with repeated measurements of exposures and covariates can be seen as a type of serial mediation model^12^. In a serial mediation model, the total effect of an exposure on the outcome can be expressed as the sum of the direct effect, and the specific indirect effects through each of the intermediate variables. We define the direct and mediated effect (DME) of an exposure at a particular timepoint as the sum of the direct effect from the exposure at that timepoint to the outcome, and the specific indirect effects through all covariates measured at later timepoints, but not including any specific indirect effects through any other exposure measurements. In terms of a causal DAG, the DME consists of the direct path from the exposure to the outcome, plus all forward paths through covariates. The DME does not include any associations between exposure and outcome that occur through back-door paths, nor any paths through the exposure at other timepoints. **Figure 2** shows those paths in the DAG that comprise the DME for each measurement of the exposure, for maternal psychopathology, SES, and offspring depressive symptoms (**Figure 1**). Consequently, DME SLCMA estimates of direct and mediated effects are summaries of the observed joint distribution are informed by a DAG, and are used to determine which life-course model best summarises the observed data in a descriptive inference.

**Figure 2.**
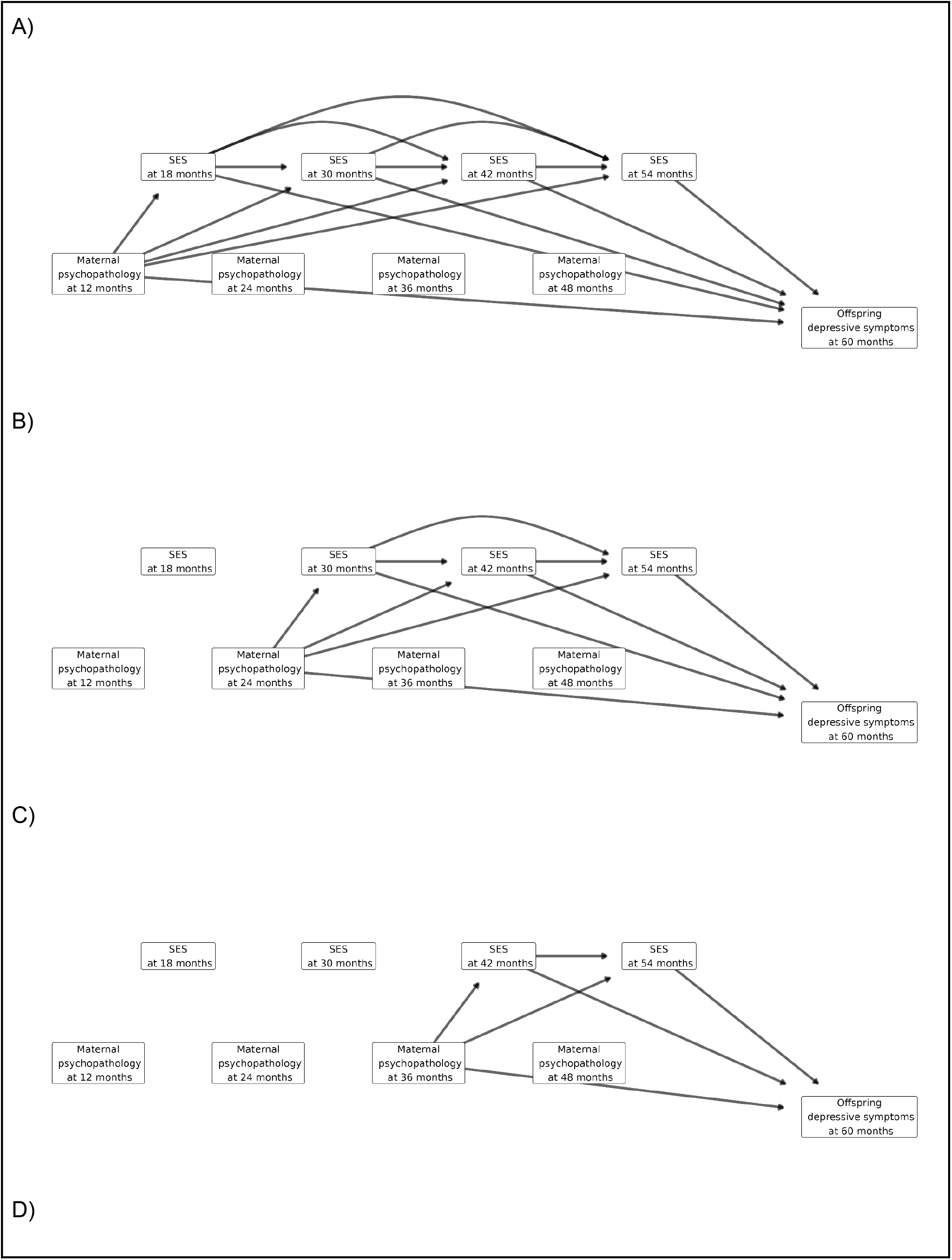
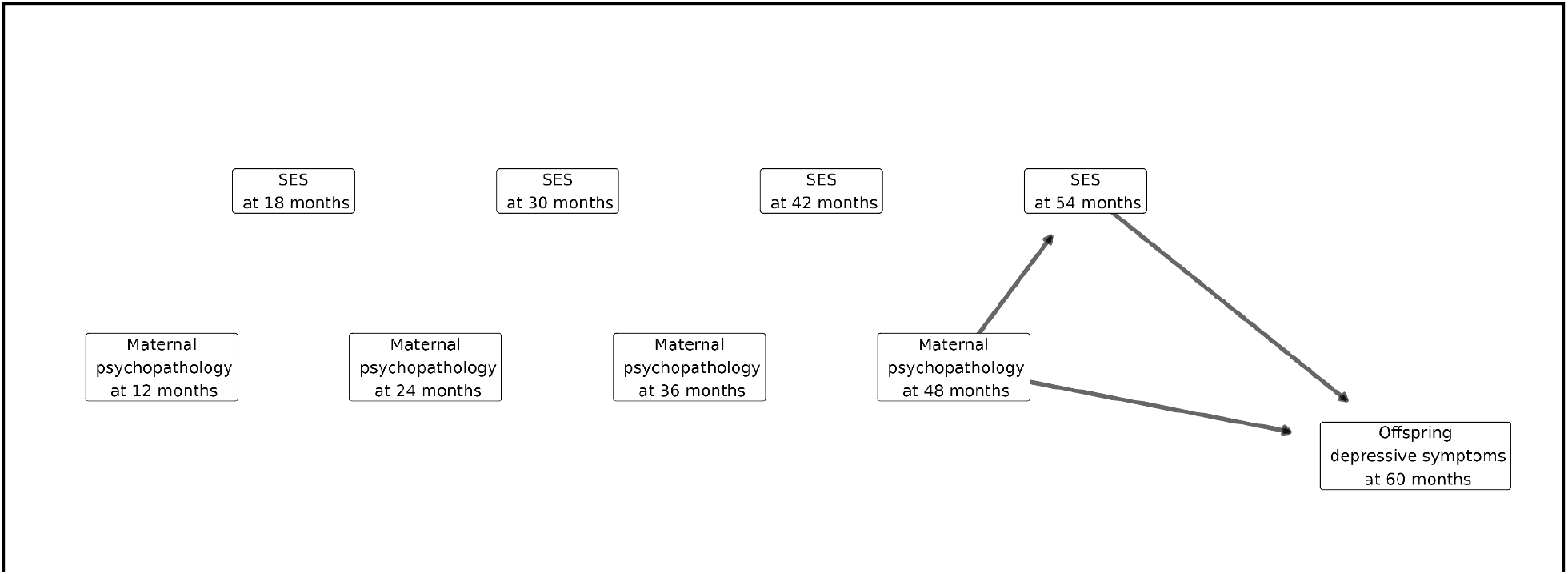
Directed acyclic graphs showing only the direct and mediated effects (DME) of maternal psychopathology on offspring depression, mediated by socioeconomic status (SES), without any back-door paths. **A)** DME of maternal psychopathology at 12 months; **B)** DME of maternal psychopathology at 24 months; **C)** DME of maternal psychopathology at 36 months; **D)** DME of maternal psychopathology at 48 months.

The DME enables the definition of life course hypotheses, analogous to the regular SLCMA, in the presence of time-varying covariates, as shown in **Table 1**.

**Table 1.**
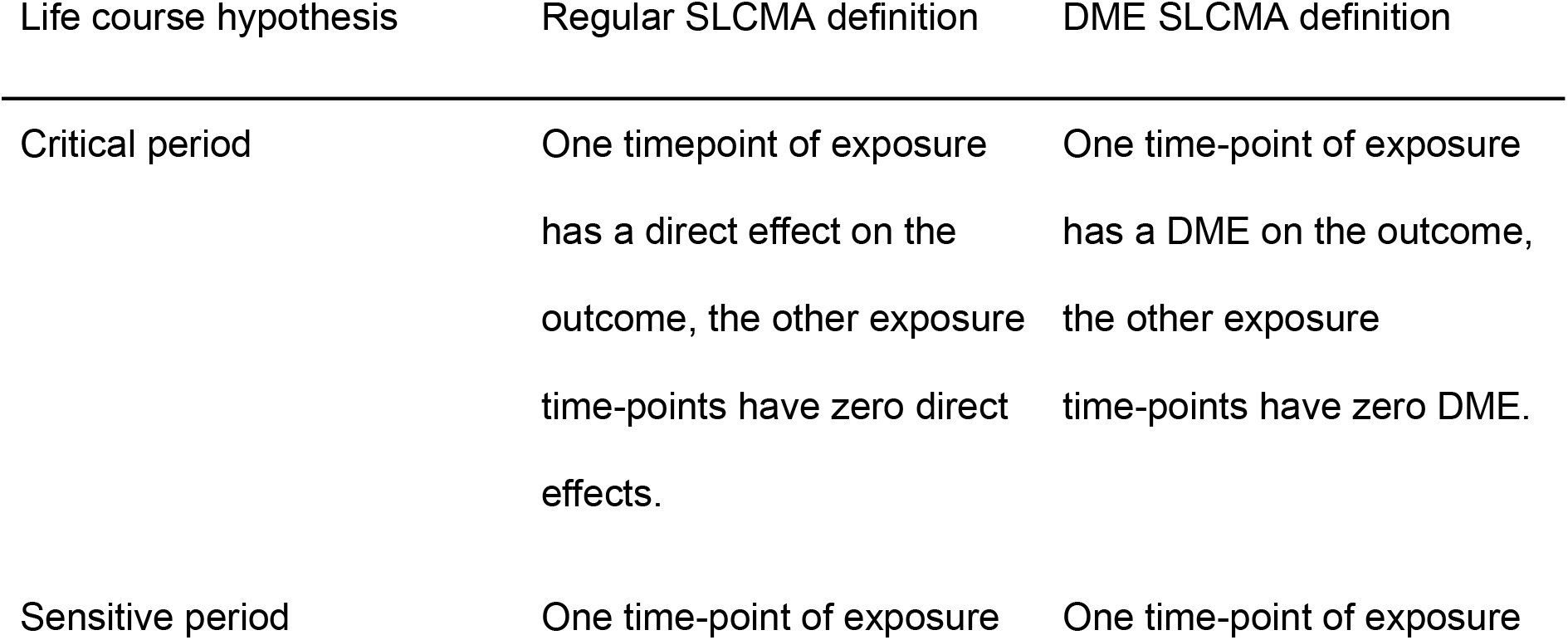
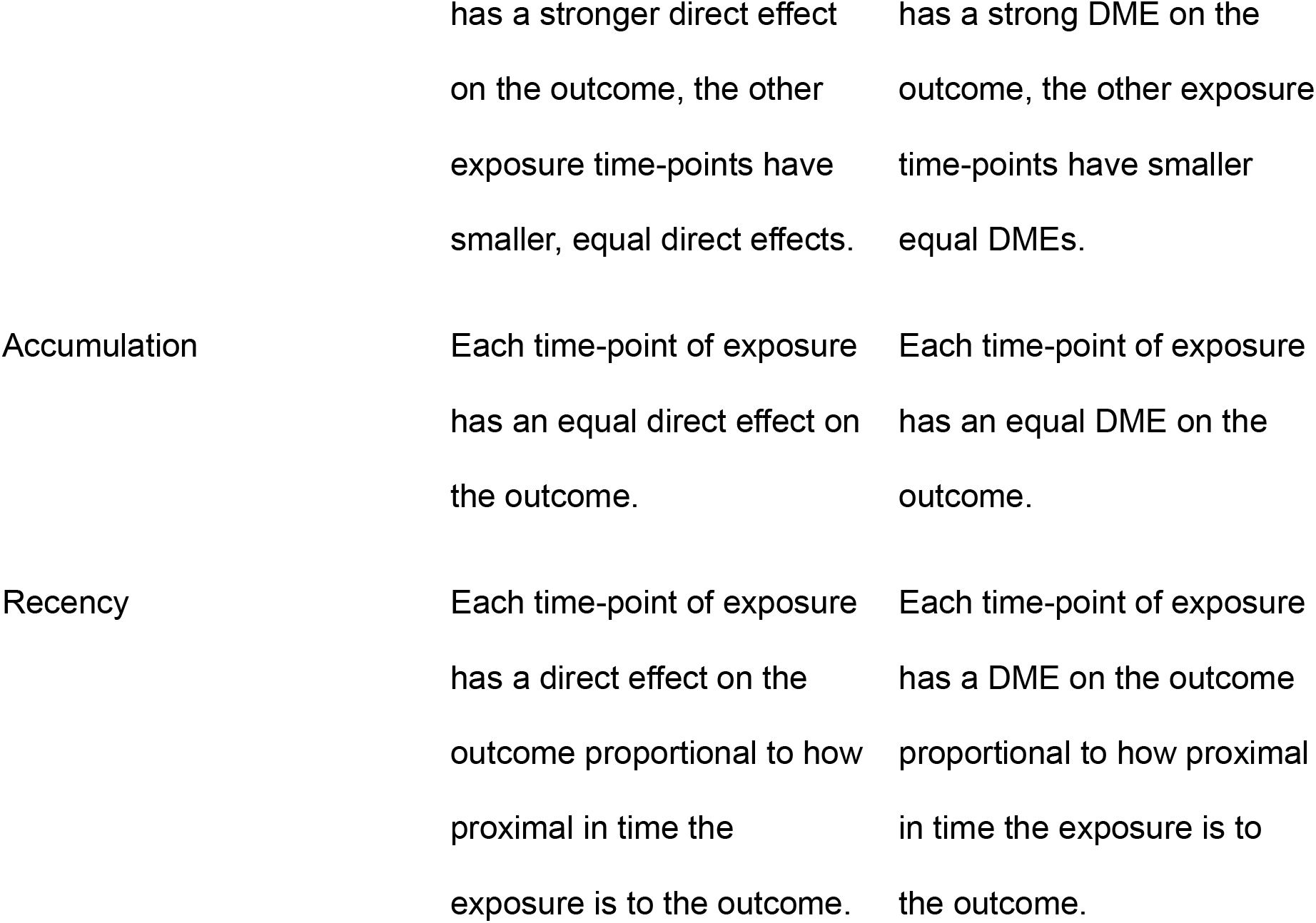
Life course hypothesis definitions. Some studies use the term sensitive period in place of critical period, while others define a sensitive period as the effect of the exposure is greater within the period while lesser, but not necessarily zero, outside the period. See paper by Schaefer et al. 2022^13^ for a synopsis of these terms as they relate to mental health.

### Estimation

In this section we present a method for constructing a consistent set of variables that can be used, in any life course regression model, to adjust for time-varying covariates and estimate appropriate DMEs.

How to estimate DMEs within a regression model without removing wanted associations, or inducing unwanted associations, requires consideration. Ignoring the covariate will allow back-door associations within the estimated effects, while adjusting for the covariate will remove back-door associations but simultaneously remove the specific indirect effects. The DME could be estimated using a different regression model (adjusting for a different set of variables) at each timepoint. However, the above models of life course hypotheses require the estimation of DMEs at all timepoints, and implementations of the SLCMA require adjusting for the same set of variables in all models. Additionally, removing the specific indirect effect through other exposure measurements presents a difficulty: adjusting for later exposure measurements leads to collider bias ^14^, inducing an unwanted association within the indirect part of the estimated effect.

Rather than adjusting for all covariates, we describe a set of residual variables such that adjusting for all residual variables results in estimation of the DMEs. This set of residual variables has a one-to-one correspondence with the set of covariate measurements. For each covariate measurement, the residual variable is obtained by regressing that variable on all preceding measurements of the exposure and covariate, then extracting the residuals from the regression. For example, for a system such as is depicted in **Figure 1** the set of residuals is:

1. The residuals from a regression of SES at 18 months on maternal psychopathology at 12 months.
2. The residuals from a regression of SES at 30 months on maternal psychopathology at 12 and 24 months, and SES at 18 months.
3. The residuals from a regression of SES at 42 months on maternal psychopathology at 12, 24 and 36 months, and SES at 18 and 30 months.
4. The residuals from a regression of SES at 54 months on maternal psychopathology at 12, 24, 36 and 48 months, and SES at 18, 30 and 42 months.

**Supplementary Information** contains a proof that this approach results in joint estimation of the DMEs in a linear model.

### Empirical example

How psychosocial factors affect childhood depression is an active area of research, including in the Drakenstein Child Health Study (DCHS) from South Africa^7,8,15^. The growing number of longitudinal psychosocial measures in such cohort studies allow for the investigation of temporal life course hypotheses. We apply DME-SLCMA to data from 336 mother child pairs in the DCHS to investigate how the timing of exposure to maternal psychopathology, from ages 12-48 months, may affect later externalising childhood depressive symptoms, at 60 months, whilst adjusting for time-varying SES. The natural logarithm of offspring depressive symptoms at 60 months is used to improve the symmetry of the outcome variable. Additional information on these variables is included in the **Supplementary Table S1**. SES would previously be adjusted for as a fixed covariate, for example using the value at birth and assuming that there is no change over time. This renders analyses prone to confounding due to any variation in SES over time and could lead to missed associations, or incorrect associations being identified, as it is plausible that SES may affect both maternal psychopathology and offspring depressive symptoms and hence confound estimates of their association.

In applying DME SLCME we first define five life course hypotheses to be investigated, selecting the single best hypothesis:

- Critical period at 12 months, critical period at 24 months, critical period at 36 months, critical period at 48 months
- Accumulation across the period 12-48 months

In this initial application of DME-SLCMA we restricted our analyses to these simple hypotheses only; it is possible to include other hypotheses, and/or compound hypotheses, in the selection process.

The critical period at 48 months was selected by DME-SLCMA as the life course hypothesis that best explains the effects of maternal psychopathology over the childhood life course on offspring depressive symptoms at 60 months of age. Post selection inference was conducted on the critical period model using the max-|*t*| test to adjust for model selection. **Table 2** shows the results of the selected model with the effect estimate values transformed back to the CBCL externalising depressive symptoms score scale from the log outcome used in analyses. We find that, on average, the depressive symptoms score at age 5 increases by a factor of 1.039 (1.010-1.069 95% CI) for each additional unit of maternal psychopathology experienced at 48 months while adjusting for time-varying SES.

**Table 2.** Empirical application results from DME SLCMA application. Max-|*t*| used in post-selection inference. *P*-value from max-|*t*| adjusts for model selection. CI has at least 95% confidence that the interval contains the population regression coefficient, adjusting for model selection. Alternative post-selection inference methods and associated results are displayed in the **Supplementary Table S4**.

|  | Estimate | Lower 95% CI | Upper 95% CI | *P*-value |
| --- | --- | --- | --- | --- |
| Critical period at<br>48 months factor | 1.039 | 1.003 | 1.076 | 0.028 |

Regular SLCMA selected the accumulation hypothesis over the critical period at 48 months due to confounding caused by the time-varying SES that could not be adjusted for. The accumulation hypothesis was the second-best hypothesis in the DME-SLCMA application - only narrowly outperformed by the critical period at 48 months. This highlights how even small amounts of confounding can lead to differences in hypothesis selections, and consequently differences in inferences made. In addition, we estimated a longitudinal causal estimate, which provides complementary evidence to our findings (and which we report in the **Supplementary Information)**.

### Simulation study theory

It is possible for the regular SLCMA to be biased by time-varying covariates, and give different conclusions from the DME-SLCMA. In this section we investigate the conditions that may cause the regular SLCMA to choose a different life course hypothesis from the DME-SLCMA due to confounding by time-varying covariates.

The impact of confounding will depend not only on the relative sizes of the effects of the exposure and covariate on the outcome, but on the timing of these effects and on the size and timing of the associations between exposure and covariate. A system with three time-points of exposure and TVC was chosen as all possible general exposure-covariate scenarios are present. We therefore conducted a simulation study (further details in **Supplementary Table S5**) in which we varied:

● **X direct effect:** the timing of the effect of the exposure on the outcome (no effect, direct effects at the first, second and third exposure time-points, accumulation)
● **C direct effect:** the timing of the direct effect of the covariate on the outcome (no effect, direct effects at the first, second and third covariate time-points, accumulation)
● **C direct effect size:** the relative size of the covariate’s direct effect on the outcome compared with the exposure’s (0.5, 1, 2, 5 times larger)
● **Correlation structure:** the structure of the correlation between exposure and covariate

`o` **Observed**: similar to that observed in our empirical example
`o` **X to C greater**: correlation arising when exposure to covariate direct effect greater than covariate to exposure direct effect
`o` **C to X greater**: correlation arising when covariate to exposure direct effect greater than exposure to covariate direct effect
`o` I**ncreasing**: correlation increases over time
`o` **Decreasing**: correlation decreases over time
● **Correlation strength:** the strength of the correlation between exposures and covariates (none, observed, twice observed, very strong)

### Deterministic simulation study

We conducted a deterministic factorial simulation study comparing the hypothesis selection between regular and DME-SLCMA. The factorial design allowed us to cover most reasonable scenarios, and was computationally feasible as deterministic simulations only require one iteration per scenario.

For each simulation, we simulated 500 measurements of maternal psychopathology across three points in time, SES at three points in time - in between each exposure measure and the following measure - and offspring depressive symptoms. The timing and size of all exposure and covariate effects and correlations were exactly controlled according to the simulation design. **Figure 3** depicts the proportion of iterations for each of the simulation scenarios in **Supplementary Table 5** where regular SLCMA disagreed on a hypothesis selection with DME-SLCMA (i.e. scenarios where regular SLCMA is confounded to a degree that causes an incorrect hypothesis selection).

**Figure 3.**
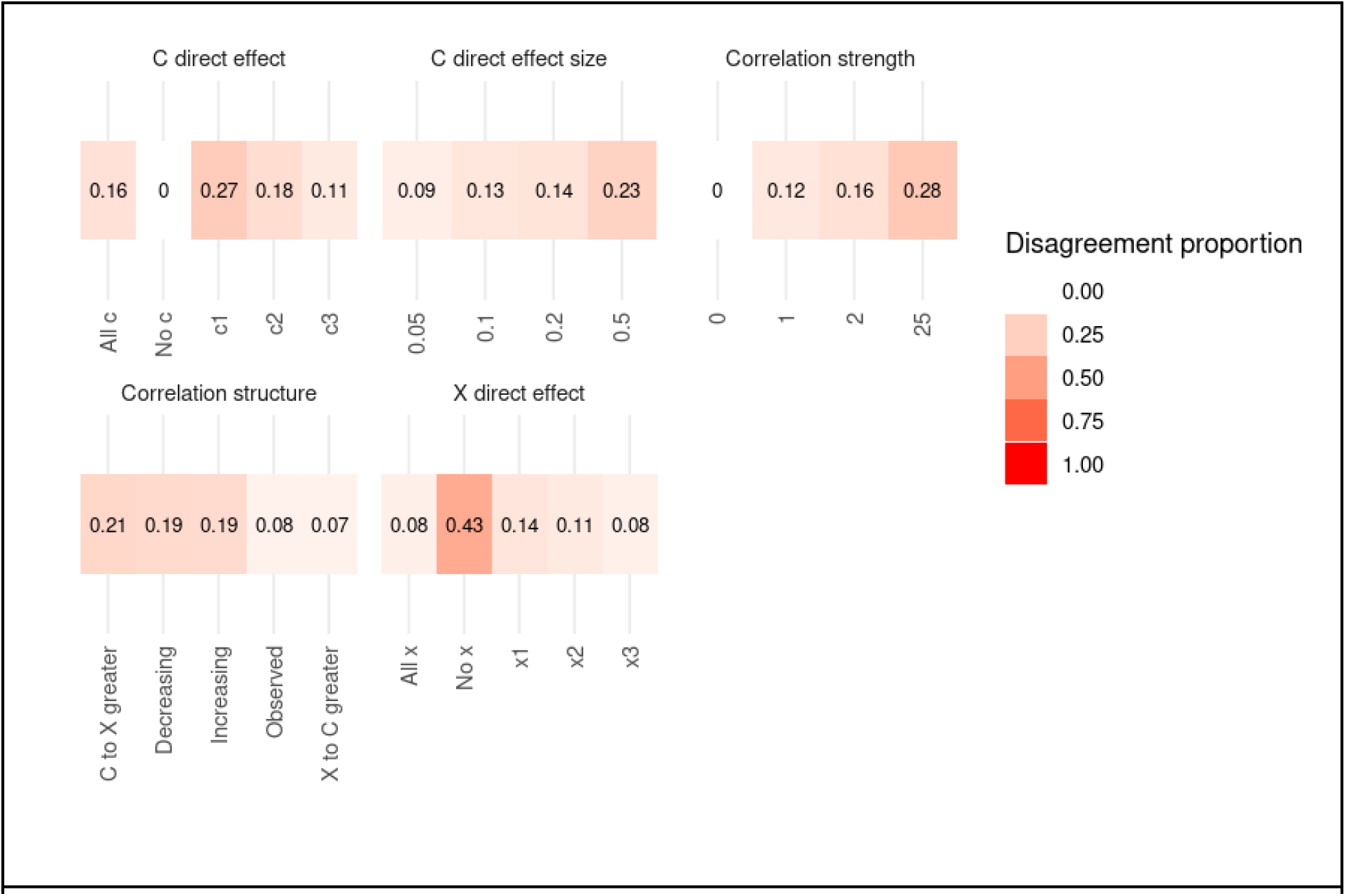
Deterministic simulation study results: Disagreement proportion across all scenarios at each level of the factors combined.

Regular SLCMA is most susceptible to selecting an incorrect hypothesis in the presence of time-varying confounding when there are no exposure direct effects present, as shown in **Figure 3**. As covariate direct effects on the outcome increase in magnitude, regular SLCMA is more likely to select an incorrect hypothesis. Additionally, regular SLCMA is more likely to select an incorrect hypothesis when the covariate direct effect on the outcome occurs earlier. As the correlation between exposures and covariates is increased, regular SLCMA selects more incorrect hypotheses. Regular SLCMA is most susceptible to confounding for systems with a covariate to exposure greater correlation structure, selecting incorrect hypotheses at an increased rate at lower levels of exposure-covariate correlation than the other correlation structures.

These deterministic simulations show the importance of adjusting for time-varying covariates to avoid incorrect hypothesis selections, especially when there are indirect effects present and no / weak direct effects from exposures. There are realistic scenarios for all correlation structures where regular SLCMA is confounded. Additional plots showing the proportion of disagreement between methods across levels of two factors are shown in **Supplementary Figures S4A-D**.

We use these deterministic simulation results to inform our selection of a baseline for stochastic simulations.

### Simulation study stochastic methods

We additionally conducted stochastic simulations for a carefully chosen number of scenarios to investigate differences between the two methods when random noise is present. To reduce the computation burden of stochastic simulations we employed a semi-factorial design with 1000 iterations for each factor level.

The exposure and covariate data for a baseline scenario, shown in **Figure 4A**, was simulated with 1000 iterations for each semi-factorial level using the same method as for the deterministic simulations. In addition to the factors used in the deterministic simulations, we introduce factors for sample size (*n = 100, 500, 1000*) and measurement error by varying *R*^2^ (0.01, 0.1) to our semi-factorial simulation design. Regular and DME-SLCMA were applied to the simulated data for each iteration in every scenario with effect estimates and hypothesis selections recorded. Proportional agreement and effect estimates between the methods for selected factors are shown in **Figures 4A-D** and all others in the **Supplementary Figures S5 and S6.**

**Figure 4.**
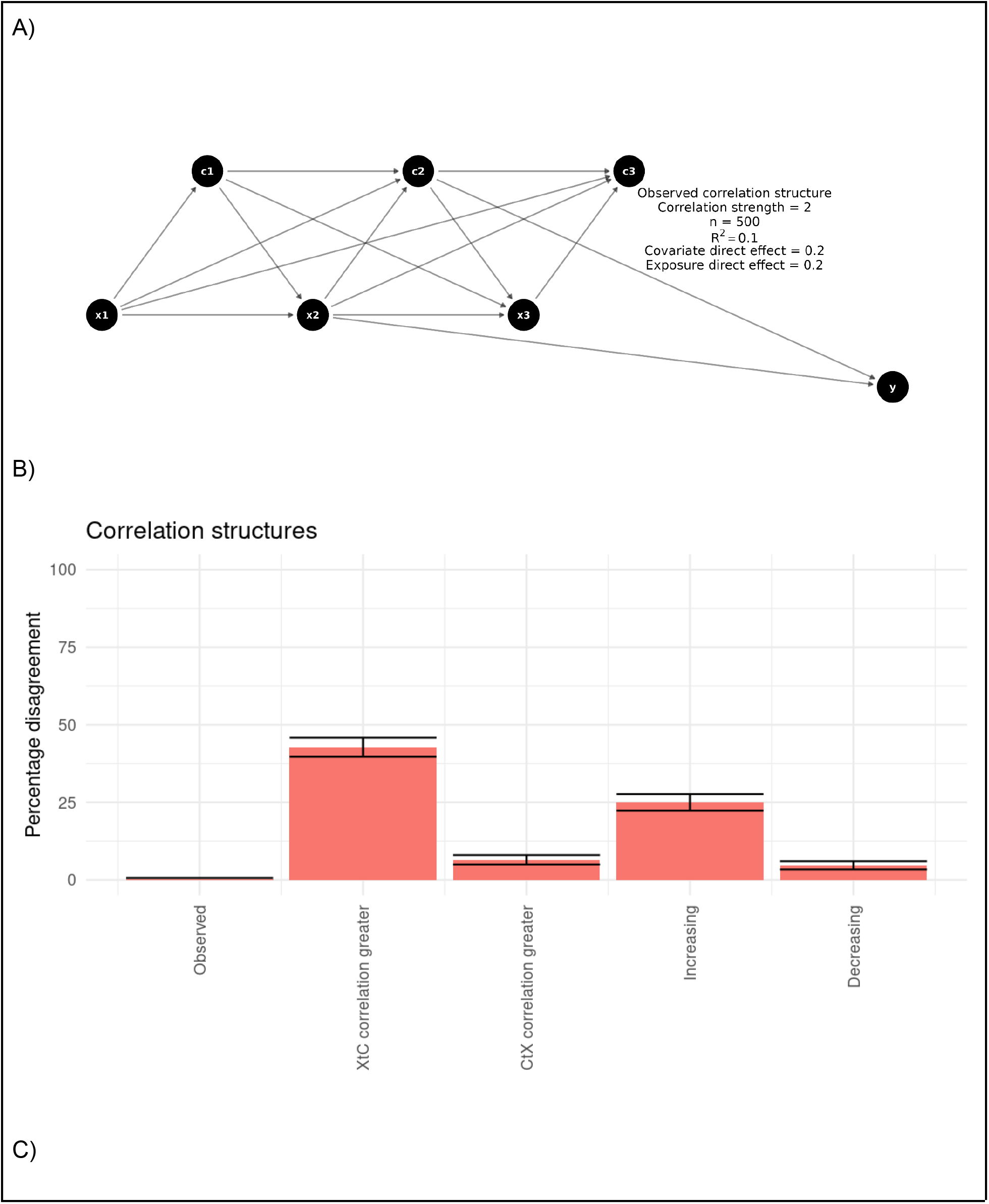
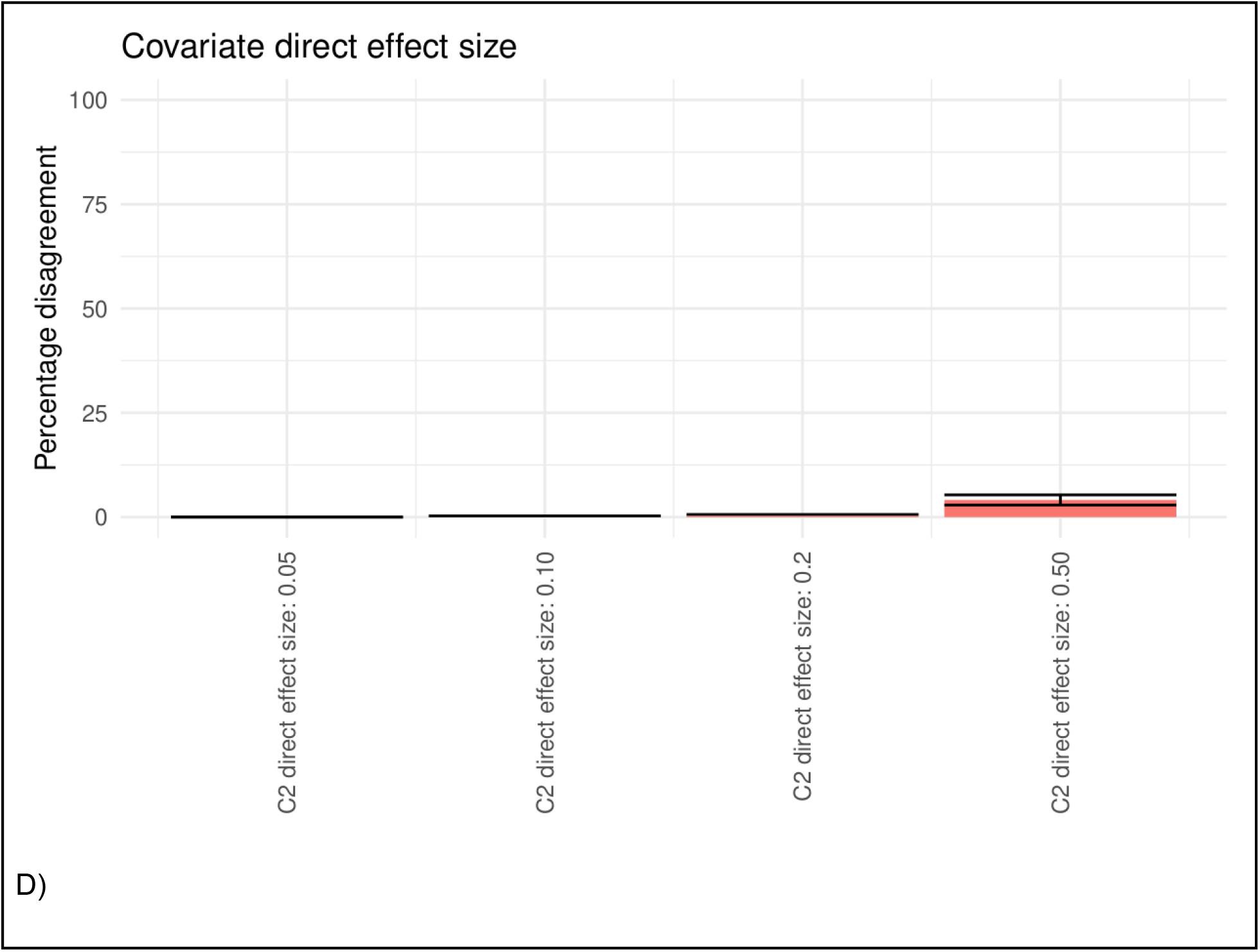
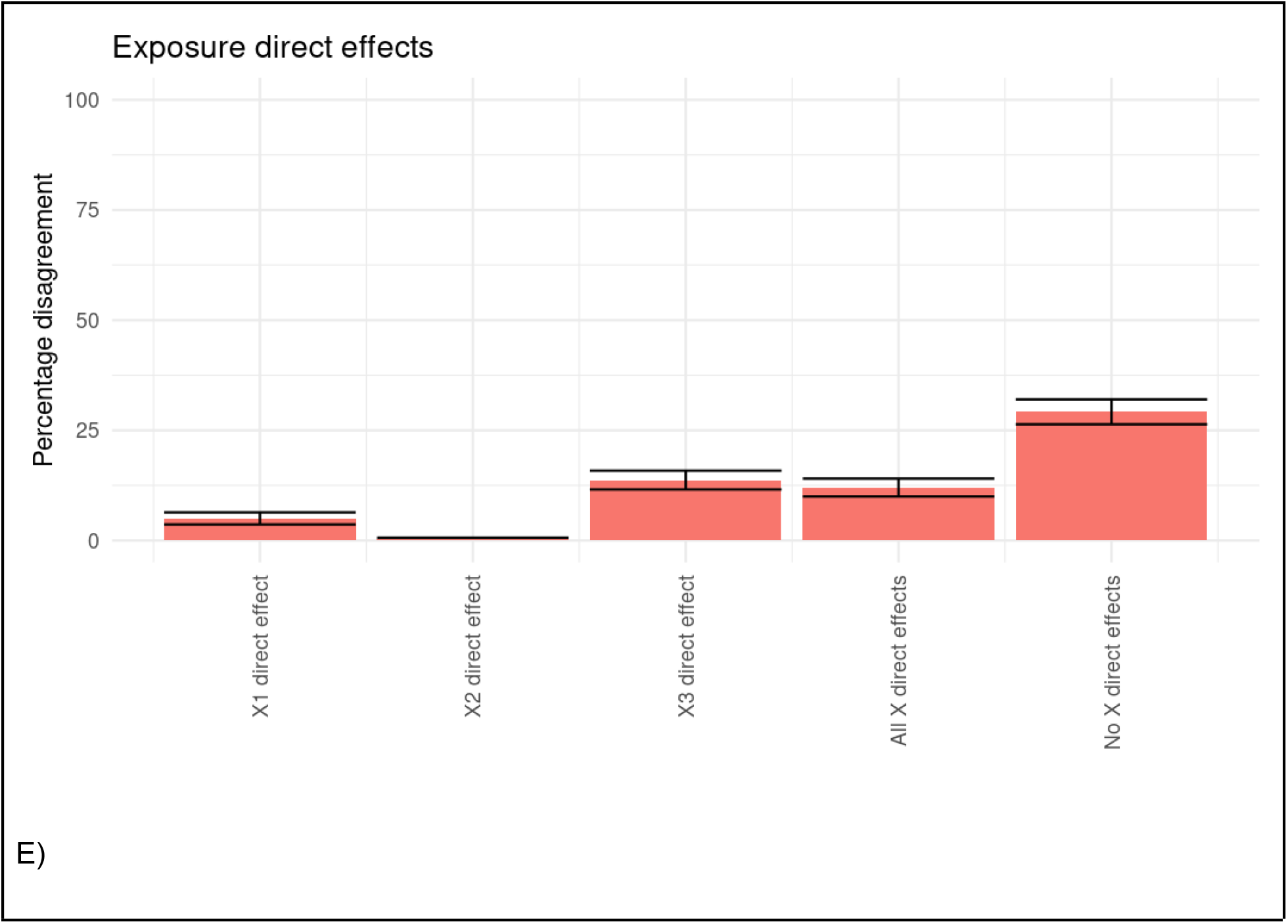
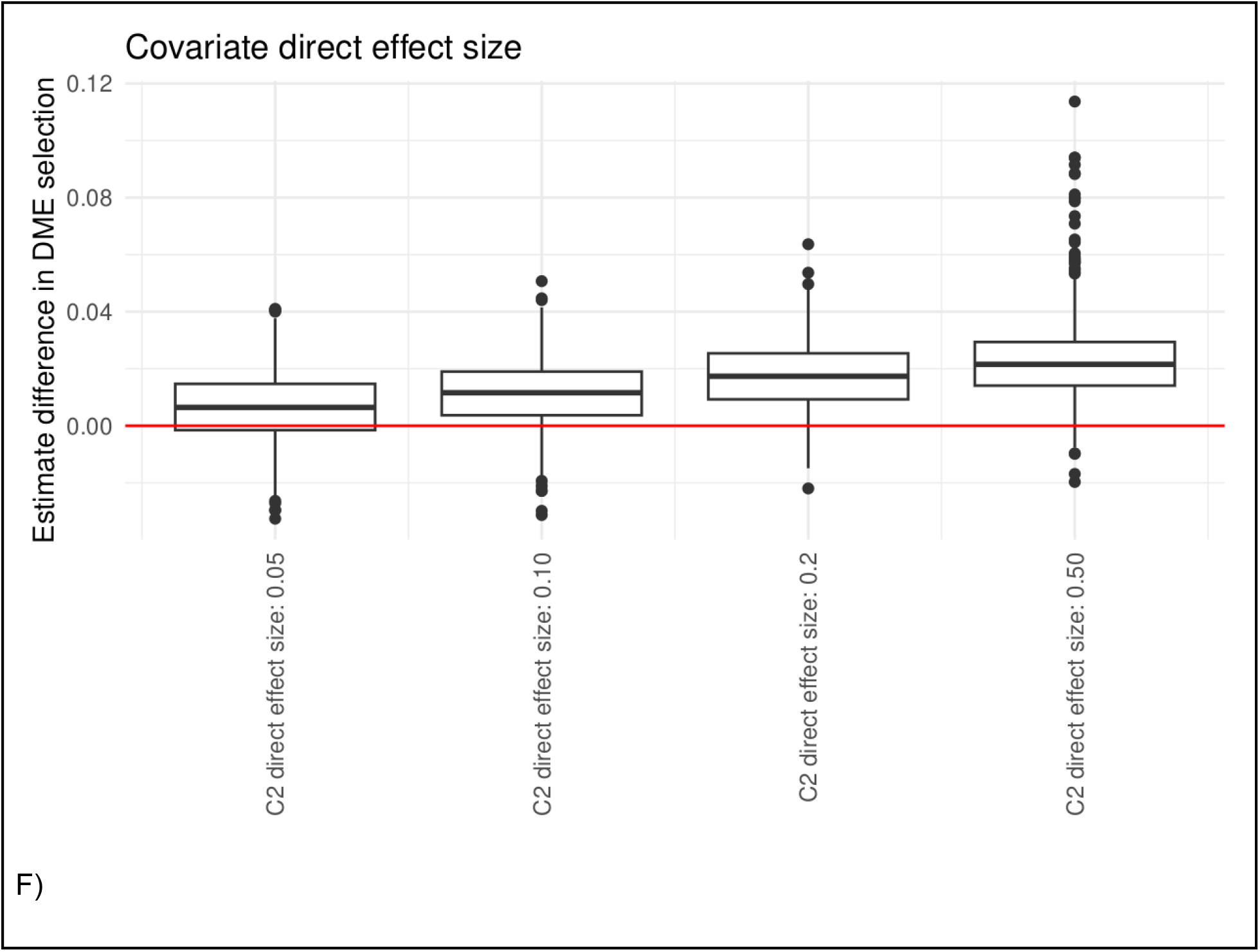
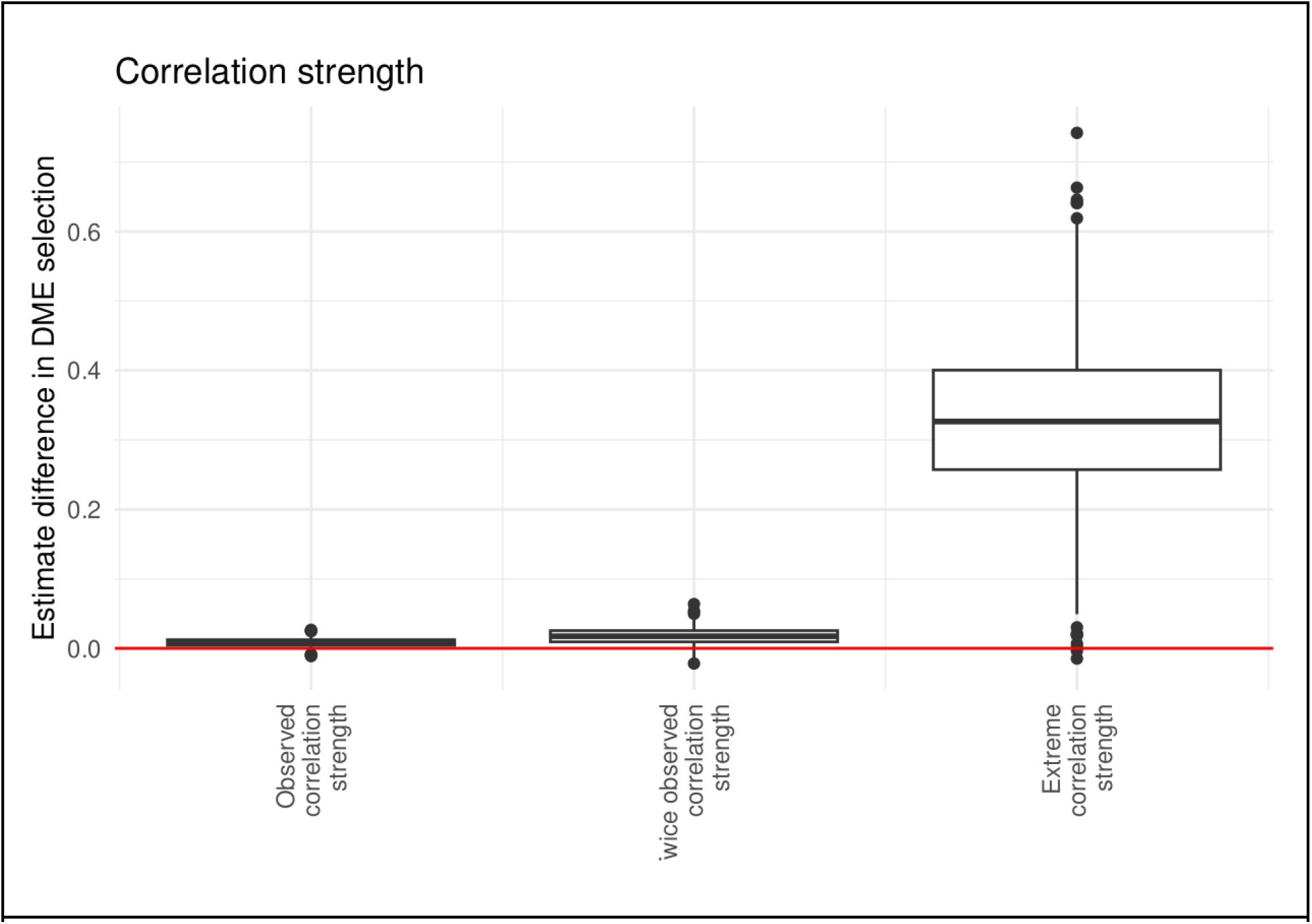
Stochastic simulation results. **A)** Stochastic simulations baseline. **B-D)** Percentage disagreement in 1000 iterations as factors are varied from baseline. **B)** Correlation structure. **C)** Covariate direct effect **D)** Exposure direct effects timing. **E)** Effect estimate bias in each iteration with increasing covariate direct effect size. **F)** Effect estimate bias in each iteration with increasing exposure-covariate correlation. Bootstrapped confidence intervals are displayed in the plots in **B-D**.

### Simulation study stochastic results

The agreement between methods on the hypothesis selected was above 90% for the baseline scenario (left column, **Figure 4B**), and most variations on this scenario. The exceptions were: X to C correlation structure, no exposure direct effects, Increasing correlation structure, a direct exposure effect from X3 and then direct exposure effects from all exposures. When there is disagreement between the methods, regular SLCMA selects a competing, but not best, hypothesis due to confounding causing bias that favours the competing hypothesis. In the baseline scenario, confounding present due to time-varying covariates is not large enough to cause regular SLCMA to select an incorrect hypothesis. When there are no exposure direct effects any confounding bias present is larger relative to the mediated effect, and regular SLCMA is more likely to select an incorrect hypothesis. Changing which exposure timepoint has a direct effect on the outcome alters the relative magnitudes of the direct and mediated effects of competing hypotheses which can increase the chance that regular SLCMA selects an incorrect hypothesis. Increasing the effect from covariates on the outcome increases both any mediated effect through that covariate, but also increases the confounding effect of that covariate and any previous covariate for which it mediates an effect on the outcome. **Figure 4E** illustrates this, with the bias due to time-varying covariates of regular SLCMA effect estimates increasing as the covariate direct effect size is increased. The same effect is observed when the exposure-covariate correlation strength is increased, which also increases both the mediation and confounding of time-varying covariates, in **Figure 4F**. Plots for the semi-factors covariate direct effects, correlation strength, sample size and noise, which each had little effect on disagreement proportions in our simulations, are shown in **Supplementary Figures S5A-D**.

## Discussion

We have shown that time-varying covariates may confound the association between an exposure and outcome and existing methods may select an incorrect life course model from this association. In response, we introduce the DME-SLCMA which selects life course hypotheses while accounting for time-varying covariates and avoiding incorrect hypothesis selections and biased results. Our method adjusts only for time-varying covariates in the past on a per hypothesis basis so that backdoor paths through covariates are blocked but all forward paths through covariates are open.

Applying the DME-SLCMA to data from the Drakenstein Child Health Study we found that a critical period at 48 months life course hypothesis for the effects of maternal psychopathology on later offspring depressive symptoms, while adjusting for time-varying socioeconomic status, best explains the variation in offspring depressive symptoms at 60 months of age. A study in the Avon Longitudinal Study of Parents And Children (ALSPAC)^17^, found an association between socioeconomic status and offspring depression which indicates that indirect effects through socioeconomic status are plausible in cohort studies. Consequently, confounding may be present if there are effects between an exposure and socioeconomic status and time-varying socioeconomic status is not adjusted for. We demonstrated that results from DME SLCMA and the marginal structural models are in accord. However, unlike DME SLCMA, there is no protocol in marginal structural models to select between life course hypotheses for that which best explains variation in the outcome. Regular SLCMA, which does not adjust for time-varying covariates, identifies an accumulation hypothesis as best, in conflict to both DME SLCMA and the marginal structural model.

In previous applications of SLCMA^18–25^ when time-varying covariates have been present, across social, environmental and economic variables, they have been considered fixed across all exposure timepoints. For example, Hardi et al. 2025^24^ adjusted for potentially time-varying child temperament measures of shyness and emotionality at age 1 as covariates fixed in time. There was only a single measure of these covariates at the time of the study, so it would not be possible to adjust for these TVC. In such cases, our simulation study results can be used to postulate the influence of confounding by unmeasured TVCs, provided that a) a later measure of the TVC were acquired and thus the correlations between each measure and both the exposures and outcome in the study could be calculated or b) the correlation structure between TVC, exposures and outcome could be hypothesized based on careful reasoning or evidence from other cohorts.

Our simulation study identifies scenarios in which confounding by a TVC is likely to result in incorrect hypothesis selections and bias if not properly adjusted for (**Table 3**). When there are only weak correlations between exposures and TVCs our simulations find that it is unlikely that regular SLCMA would select an incorrect hypothesis. However, if there are two closely competing life course hypotheses, any small level of TVC confounding can cause regular SLCMA to select an incorrect hypothesis. We recommend using DME-SLCMA to adjust for TVC if, in a particular application, a TVC of confounding concern is identified – one that fulfills the conditions in **Table 3**.

**Table 3.** Scenarios where estimates obtained ignoring time-varying covariates are most susceptible to incorrect hypothesis selections due to confounding.

Selections made ignoring time-varying covariates are likely to be confounded when all the following conditions are met:

- There are no, or only weak, direct effects from exposures on the outcome.
- The exposures of interest are likely to have mediated effects on the outcome.
- There are strong exposure-covariate correlations, as well as covariate-outcome correlations

Our method provides unbiased life course hypothesis selection and effect estimation in the presence of time-varying covariates and indirect effects and is highly computationally efficient. Using the marginal structural models (see **Supplementary Information** for details), we found agreement between this method and DME SLCMA: the marginal structural model estimated the effect of binary exposure to maternal psychopathology at 48 months of age to have the greatest effect on offspring depression at 60 months of age, as shown in **Table S2**. To compare between an accumulation and 48 month critical period hypothesis we implemented an alternative marginal structural model, **Model S2**, and found evidence to support the 48 month critical period hypothesis over an accumulation hypothesis. These results suggest that to reduce depression in offspring at 60 months, an intervention on maternal psychopathology at 48 months could be efficacious. However, data on the magnitude of a specific therapeutic intervention would be required to estimate the effect of a possible intervention. Practically, the life course hypothesis selected by DME SLCMA, that which best explains the observed data, is suitable and in agreement with the causal effects from the marginal structural model.

A limitation of our method is that it relies on the correct specification of a DAG when encoding each life course hypothesis and its relationship with the covariate. In our empirical example, the longitudinal spacing of exposures and covariates made it easier to infer the direction of causal effects. This is more difficult in studies where the exposure and covariate may be measured at the same time. Additionally, we assumed negligible effects of any other potentially confounding time-varying covariates, such as time-varying childhood behaviour, which may affect offspring depression.

## Data Availability

An anonymised, de-identified version of the dataset can be made available on request. All requests should be directed to Heather Zar, the Principal Investigator of DCHS.

## Funding

This work was supported by the National Institute of Mental Health of the National Institutes of Health [R01 MH113930; PI: Dunn] and Research Ireland through the Research Ireland Centre for Research Training in Genomics Data Science [18/CRT/6214]. The NIMH had no further role in study design; in the collection, analysis and interpretation of data; in the writing of the report; and in the decision to submit the paper for publication. The content is solely the responsibility of the authors and does not necessarily represent the official views of the National Institutes of Health. The Drakenstein Child Health Study (DCHS) was supported by the Bill and Melinda Gates Foundation [OPP1017641,OPP1017579]; the National Institute of Mental Health [1R21MH098662–01]; the National Institute of Health H3Africa [1U01AI110466-01A1]; the National Research Foundation; the South African Medical Research Council and the Wellcome Trust [221372/Z/20/Z].

## Acknowledgements

We greatly thank the parents and children who participated in this study. We would also like to thank the study staff in Paarl, the study data and laboratory teams, and the clinical and administrative staff of the Western Cape Government Health Department at Paarl Hospital and at the clinics for support of the study.

## Author contributions

ADACS developed the novel method, conducted the literature review and helped prepare the Introduction and Methods section of the text. ADACS, AJS and ECD directed the study’s implementation. SB conducted the simulation study, empirical application and drafted the manuscript. SYE helped implement the empirical application. NFS helped implement and prepare the Empirical Applications section of the text. HJZ is the principal investigator of the DCHS who conceived and designed the cohort study and obtained core funding, together with DJS who is the psychosocial lead of the DCHS. All authors contributed to the final manuscript.

## Supplementary data

Supplementary data is made available online.

## Conflict of interest

None declared.

## Notes

### Competing Interest Statement

The authors have declared no competing interest.

### Author Declarations

Human Research Ethics Committee, Faculty of Health Sciences, University of Cape Town (401/2009) and Stellenbosch University (N12/02/2009) gave ethical approval for this work. The Western Cape Provincial Health Research committee (2011RP45) gave ethical approval for this work.

### Summary of Updates

Discussion updated in include complementary causal methods. Supplemental files updated. Author list updated.

