## Supplementary Information for "Direct and mediated effects (DME) SLCMA: a novel method for life course modelling with time-varying covariates"

### Supplementary material

#### Full proof that adjusting for residual variables results in joint estimation of the DMEs

In this section we consider a general situation of an arbitrary number of repeated exposures and an arbitrary number of repeated covariates.

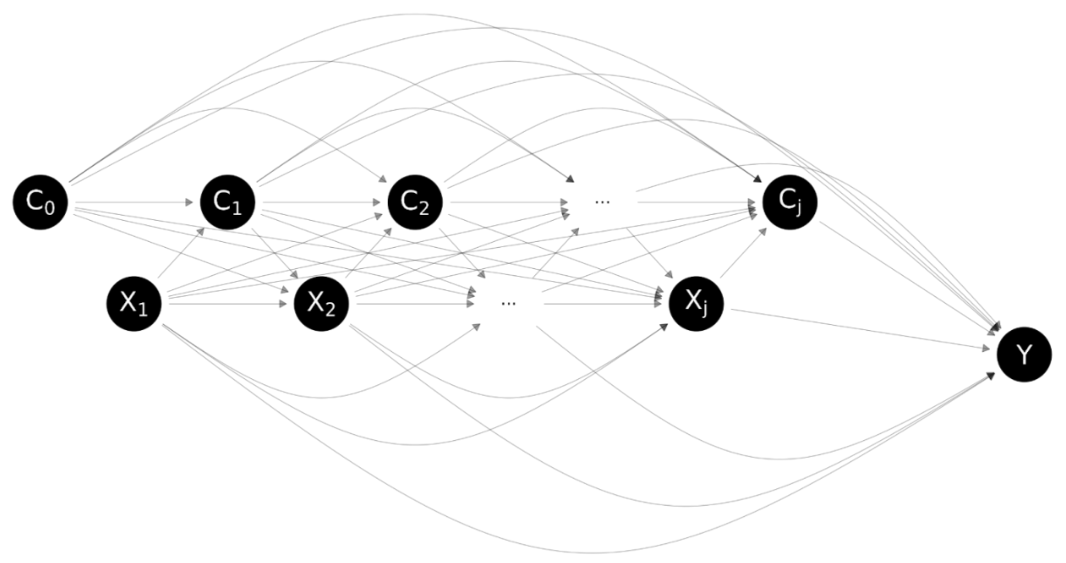

**Figure S1.** A general causal DAG for a life course modelling scenario. Measures of a covariate over time are represented by $C_i$, the exposure over time by $X_0$ and the later outcome by $Y$.

**Figure S1** depicts a general scenario with $J$ repeated exposures $X_1, X_2, \dots, X_J$ and a later outcome $Y$. In addition, there may be a baseline covariate ($C_0$), which precedes all exposures in the causal DAG, and $J$ subsequent covariates $C_1, C_2, \dots, C_J$. We assume that the timepoints for exposures and covariates alternate through the lifecourse, so that $C_{j-1}$ precedes $X_j$, and $X_j$ precedes $C_j$, for all $j = 1, \dots, J$. The DAG in Figure S1 does not show all possible causal pathways for the sake of legibility. For full generalisability we consider that every exposure $X_1, X_2, \dots, X_J$ may have a direct effect on $Y$, every covariate $C_0, C_1, \dots, C_J$ may have a direct effect on $Y$, every exposure may have a direct effect on every subsequent covariate (i.e., $X_j$ may have a direct effect on $C_j, C_{j+1}, \dots, C_J$ for all $j = 1, \dots, J$), and every covariate may have a direct effect on every a subsequent exposure (i.e., $C_{j-1}$ may have a direct effect on $X_j, X_{j+1}, \dots, X_J$ for all $j = 1, \dots, J$). We also allow for multiple covariates at each timepoint, so $C_j$ could represent a matrix of several covariates. We allow for multiple covariates for two reasons: i) to allow for multiple mediation with several mediating covariates, and ii) to allow for a single covariate to be repeated several times between exposure timepoints. This second reason allows generalisability beyond our assumption that the timepoints for exposures and covariates alternate through the life course.

We will define a residual variable $R_j$, for $j = 1, \dots, J$, as the residual obtained from a linear regression of $C_j$ on all previous exposures and covariates. This linear regression will use the model matrix

$$B = \begin{pmatrix} C_0 & X_1 & C_1 & X_2 & \cdots & C_{j-1} & X_j \end{pmatrix}.$$

We have used the notation $B$ to indicate the model matrix consisting of all variables that occur *before* the covariate $C_j$. Let $\alpha$ denote the coefficients in the regression of $C_j$ on the columns of $B$. Thus $\alpha$ contains the first part of the indirect effect (i.e., the exposure to covariate coefficient) through $C_j$, for every preceding exposure. We have

$$\hat{\alpha} = (B^T B)^{-1} B^T C_j.$$

Hence the residual variable may be written as

$$R_j = C_j - B(B^T B)^{-1} B^T C_j = HC_j,$$

where $H = I - B(B^T B)^{-1} B^T$. Note that the matrix $H$ is symmetrical (because $H^T = H$) and idempotent (because $H^2 = H$).

We also introduce a second model matrix

$$A = \begin{pmatrix} X_{j+1} & R_{j+1} & \cdots & X_J & R_J \end{pmatrix}.$$

We have used the notation $A$ to indicate the model matrix consisting of all exposures and residual variables that occur *after* the covariate $C_j$.

#### Lemma: effect on regression coefficients of swapping one covariate for its residual

Consider the following two linear regression models.

Model 1 has dependent variable $Y$ and predictors $X_1, X_2, \dots, X_J, C_0, C_1, \dots, C_{j-1}, C_j, R_{j+1}, \dots, R_J$ for some $j = 1, \dots, J$. That is, regression of $Y$ on all exposures, all covariates that precede $C_j$, the covariate $C_j$, and residual variables for all covariates that follow $C_j$. This linear regression will use the model matrix $X = \begin{pmatrix} B & C_j & A \end{pmatrix}$. Let $\gamma_B, \beta, \gamma_A$ denote the coefficients to be estimated by Model 1 corresponding to the columns of $B, C_j, A$ respectively.

Model 2 has dependent variable $Y$ and predictors $X_1, X_2, \dots, X_J, C_0, C_1, \dots, C_{j-1}, R_j, R_{j+1}, \dots, R_J$ for some $j = 1, \dots, J$. The predictors of Model 2 are almost identical to the predictors of Model 1, expect that the covariate $C_j$ has been swapped for the residual variable $R_j$. This linear regression will use the model matrix $X' = \begin{pmatrix} B & R_j & A \end{pmatrix}$. Let $\gamma_B', \beta', \gamma_A'$ denote the coefficients to be estimated by Model 2 corresponding to the columns of $B, R_j, A$ respectively.

The following relationships hold between the estimated coefficients of Model 1 and the estimated coefficients of Model 2:

$$\begin{aligned} \widehat{\gamma_B'} &= \hat{\alpha}\hat{\beta} + \widehat{\gamma_B}, \\ \widehat{\beta'} &= \hat{\beta}, \\ \widehat{\gamma_A'} &= \widehat{\gamma_A}. \end{aligned}$$

*Proof*

To find an expression for the estimates in Model 1, we must invert the matrix

$$X^TX = \begin{pmatrix} B^TB & B^TC_j & B^TA \\ C_j^TB & C_j^TC_j & C_j^TA \\ A^TB & A^TC_j & A^TA \end{pmatrix}.$$

The matrix inverse can be written in terms of the matrix

$$F = \begin{pmatrix} C_j^TC_j & C_j^TA \\ A^TC_j & A^TA \end{pmatrix} - \begin{pmatrix} C_j^TB \\ A^TB \end{pmatrix}(B^TB)^{-1}\begin{pmatrix} B^TC_j & B^TA \end{pmatrix}$$
$$= \begin{pmatrix} C_j^THC_j & C_j^THA \\ A^THC_j & A^THA \end{pmatrix},$$

so that the matrix inverse is

$$(X^TX)^{-1}$$
$$= \begin{pmatrix} (B^TB)^{-1} + (B^TB)^{-1}\begin{pmatrix} B^TC_j & B^TA \end{pmatrix}F^{-1}\begin{pmatrix} C_j^TB \\ A^TB \end{pmatrix}(B^TB)^{-1} & -(B^TB)^{-1}\begin{pmatrix} B^TC_j & B^TA \end{pmatrix}F^{-1} \\ -F^{-1}\begin{pmatrix} C_j^TB \\ A^TB \end{pmatrix}(B^TB)^{-1} & F^{-1} \end{pmatrix}.$$

Finally, the estimated regression coefficients are

$$\begin{pmatrix} \widehat{\gamma_B} \\ \hat{\beta} \\ \widehat{\gamma_A} \end{pmatrix} = (X^TX)^{-1}X^TY = (X^TX)^{-1}\begin{pmatrix} B^TY \\ C_j^TY \\ A^TY \end{pmatrix}$$
$$= \begin{pmatrix} (B^TB)^{-1}B^TY - (B^TB)^{-1}\begin{pmatrix} B^TC_j & B^TA \end{pmatrix}F^{-1}\begin{pmatrix} C_j^TY - C_j^TB(B^TB)^{-1}B^TY \\ A^TY - A^TB(B^TB)^{-1}B^TY \end{pmatrix} \\ F^{-1}\begin{pmatrix} C_j^TY - C_j^TB(B^TB)^{-1}B^TY \\ A^TY - A^TB(B^TB)^{-1}B^TY \end{pmatrix} \end{pmatrix}$$
$$= \begin{pmatrix} (B^TB)^{-1}B^TY - (B^TB)^{-1}\begin{pmatrix} B^TC_j & B^TA \end{pmatrix}F^{-1}\begin{pmatrix} C_j^THY \\ A^THY \end{pmatrix} \\ F^{-1}\begin{pmatrix} C_j^THY \\ A^THY \end{pmatrix} \end{pmatrix}.$$

In Model 2, note that the model matrix can be written as $X' = \begin{pmatrix} B & R_j & A \end{pmatrix} = \begin{pmatrix} B & HC_j & A \end{pmatrix}$. Further note that $B^THC_j = 0$. To find an expression for the estimates in Model 2, we must invert the matrix

$$X'^TX' = \begin{pmatrix} B^TB & 0 & B^TA \\ 0 & C_j^THC_j & C_j^THA \\ A^TB & A^THC_j & A^TA \end{pmatrix}.$$

The matrix inverse can be written in terms of the matrix

$$\begin{pmatrix} C_j^THC_j & C_j^THA \\ A^THC_j & A^TA \end{pmatrix} - \begin{pmatrix} 0 \\ A^TB \end{pmatrix}(B^TB)^{-1}(0 \quad B^TA)$$
$$= \begin{pmatrix} C_j^THC_j & C_j^THA \\ A^THC_j & A^THA \end{pmatrix} = F,$$

so that the matrix inverse is

$$\left(X'^T X'\right)^{-1} = \begin{pmatrix} (B^T B)^{-1} + (B^T B)^{-1}(0 \quad B^T A)F^{-1}\begin{pmatrix} 0 \\ A^T B \end{pmatrix}(B^T B)^{-1} & -(B^T B)^{-1}(0 \quad B^T A)F^{-1} \\ -F^{-1}\begin{pmatrix} 0 \\ A^T B \end{pmatrix}(B^T B)^{-1} & F^{-1} \end{pmatrix}.$$

Finally, the estimated regression coefficients are

$$\begin{pmatrix} \widehat{\gamma_B'} \\ \widehat{\beta'} \\ \widehat{\gamma_A'} \end{pmatrix} = \left(X'^T X'\right)^{-1} X'^T Y = \left(X'^T X'\right)^{-1} \begin{pmatrix} B^T Y \\ C_j^T HY \\ A^T Y \end{pmatrix}$$

$$= \begin{pmatrix} (B^T B)^{-1} B^T Y - (B^T B)^{-1}(0 \quad B^T A)F^{-1}\begin{pmatrix} C_j^T Y - 0 \\ A^T Y - A^T B (B^T B)^{-1} B^T Y \end{pmatrix} \\ F^{-1}\begin{pmatrix} C_j^T HY - 0 \\ A^T Y - A^T B (B^T B)^{-1} B^T Y \end{pmatrix} \end{pmatrix}$$

$$= \begin{pmatrix} (B^T B)^{-1} B^T Y - (B^T B)^{-1}(0 \quad B^T A)F^{-1}\begin{pmatrix} C_j^T HY \\ A^T HY \end{pmatrix} \\ F^{-1}\begin{pmatrix} C_j^T HY \\ A^T HY \end{pmatrix} \end{pmatrix}.$$

We can see that $\beta' = \beta$ and $\widehat{\gamma_A'} = \widehat{\gamma_A}$. We also note that

$$\begin{aligned} \widehat{\gamma_B'} - \widehat{\gamma_B} &= (B^T B)^{-1}(B^T C_j \quad 0)F^{-1}\begin{pmatrix} C_j^T HY \\ A^T HY \end{pmatrix} \\ &= (B^T B)^{-1}(B^T C_j \quad 0)\begin{pmatrix} \hat{\beta} \\ \widehat{\gamma_A} \end{pmatrix} = (B^T B)^{-1} B^T C_j \, \hat{\beta} = \hat{\alpha}\hat{\beta} \end{aligned}$$

as required. This proof assumes a linear and additive system, such that mediated effect through a pathway can be calculated by multiplying the respective path coefficients together, and the total effect the sum of all open pathways.

#### Theorem: adjusting for residual variables results in joint estimation of the DMEs

In the linear regression model with dependent variable $Y$ and predictors $X_1, X_2, \dots, X_J, C_0, R_1, \dots, R_J$, the estimated coefficients for $X_1, X_2, \dots, X_J$ are the direct and mediated effects of $X_1, X_2, \dots, X_J$, respectively, on $Y$.

##### *Proof*

This can be seen by starting with the linear regression model with dependent variable $Y$ and predictors $X_1, X_2, \dots, X_J, C_0, C_1, \dots, C_J$. In this starting model, the estimated coefficients for $X_1, X_2, \dots, X_J$ consist of the direct effects of these exposures on the outcome, with no bias due to confounding as all covariates are adjusted for in the model. From this starting model, we repeatedly applying the above Lemma, each time swapping a covariate $C_j$ is swapped for a residual variable $R_j$ (starting with $C_J$). As proved in the above Lemma, this swap does not alter the regression coefficient for any exposure that follows $C_j$ (so the swap does not introduce confounding bias). Also proved in the above Lemma is this swap adds the indirect effect through $C_j$ to the regression coefficient for all exposures that precede $C_j$. Once all covariates $C_1, \dots C_J$ have been swapped for residual variables, the overall result is that the regression coefficient for every exposure consists of the direct effect plus the indirect effect through all following covariates, while adjusting for confounding by all preceding covariates.

#### Empirical example

Data were obtained from the Drakenstein Child Health Study (DCHS), a multidisciplinary longitudinal birth cohort from the Western Cape of South Africa[1,2]. The study was approved by the faculty of Health Sciences, Human Research Ethics Committee, University of Cape Town (401/2009), Stellenbosch University (N12/02/0002) and the Western Cape Provincial Health Research committee (2011RP45). Pregnant mothers were enrolled at 20-28 weeks' gestation from two clinics, Mbekweni and TC Newman, during routine antenatal care appointments between March 2012 and March 2015. The Mbekweni clinic predominantly serves an isiXhosa-speaking Black African population, whereas the TC Newman clinic predominantly serves an Afrikaans-speaking mixed ancestry population.

In South Africa, the apartheid system divided individuals into different racial groups (e.g., "Black African", "Coloured", "White"). Under democracy these categories continue to be employed, partly for reasons of redress and partly due to South Africans' cultural embrace and celebration of these identities[3]. In DCHS, participants self-identify as "Black African" or "Mixed ancestry". We are aware that these categories are not widely used internationally and may have painful racist connotations in other cultures. At the same time, we think it is important to honour South African participants' identities and culture by using the language and terminology they self-report. By using these categories, we are not aiming to reify social categories but instead hope to contribute to the study of ongoing health disparities stemming from systemic racism.

Both clinics are located in the peri-urban Drakenstein subdistrict. To be eligible for the study, women needed to: (1) be 18 years of age or older, (2) intend to attend antenatal care at one of the two clinics, and (3) intend to remain in the area for at least a year. Participants provided informed consent in their preferred language (English, isiXhosa, or Afrikaans), which was renewed annually. Ethics approval was provided by the Human Research Ethics Committee of the Faculty of Health Sciences at the University of Cape Town, Stellenbosch University, and the Western Cape Provincial Research committee.

A total of 1225 mothers were originally enrolled in the study, with 1137 giving live birth to 1143 children. The remainder of mothers either experienced stillbirth (n=13, two sets of twins), miscarriage (n=9), or were lost to follow-up prior to their delivery (n=67). Four mothers had twins and one had triplets; these non-singleton births were removed from analyses.

For our analyses, the exposure, maternal psychological distress, was measured at 12 months, 24 months, 36 months and 48 months using the Self-Reporting Questionnaire-20 (SRQ-20)[4]. Each item was coded based on whether the symptom was present (1) or absent (0) at the time of collection. Individual items were summed to generate a total score, with higher scores indicating greater psychological distress, including depression and anxiety. The time-varying socioeconomic status was measured using a standardised score summing four socioeconomic components: education, income, assets and employment. The education component measures the level of caregiver education obtained, with three levels, primary (0), some secondary (1) and complete secondary (2). The average household income over the past 6 months is stratified into <R1000 a month (0), R1000-5000 a month (1), and >R5000 a month (2). Assets sum (0-13) was measured from a yes (1), no (1) answers in a questionnaire to the presence of the following in the household: Electricity, running water, domestic worker, flush toilet inside, built in kitchen sink, electric stove/hotplate, working phone, at least one motor vehicle, motorcycle/scooter, bicycle, shop at supermarkets, use of financial services, account at a retail store. Caregiver employment was measured as not-working (0) and working (1). These four components were summed and standardised to give the socioeconomic measure for each of the time-points. This is a socioeconomic measure designed for use in the DCHS, and has been adapted from that used in the South African Stress and Health Study[5]. The outcome, offspring depressive symptoms, was measured at 60 months using the externalising problems subscore of the Child Behavioural CheckList (CBCL)[6,7], taking the natural log to improve outcome symmetry.  The 113-item scale references behaviours over the last 6 months, with responses ranging from 0 (“Not true (as far as you know)”) to 2 (“Very true or often true”). The CBCL includes nine subscales: anxiety, depression, somatic complaints, social problems, thought problems, attention problems, rule-breaking behaviour, aggressive behaviour, and other problems (such as problems with sleep). These subscales can be combined to form internalizing and externalizing subscales. The 336 individuals with complete data of these variables and timepoints were used in this analysis.

A)

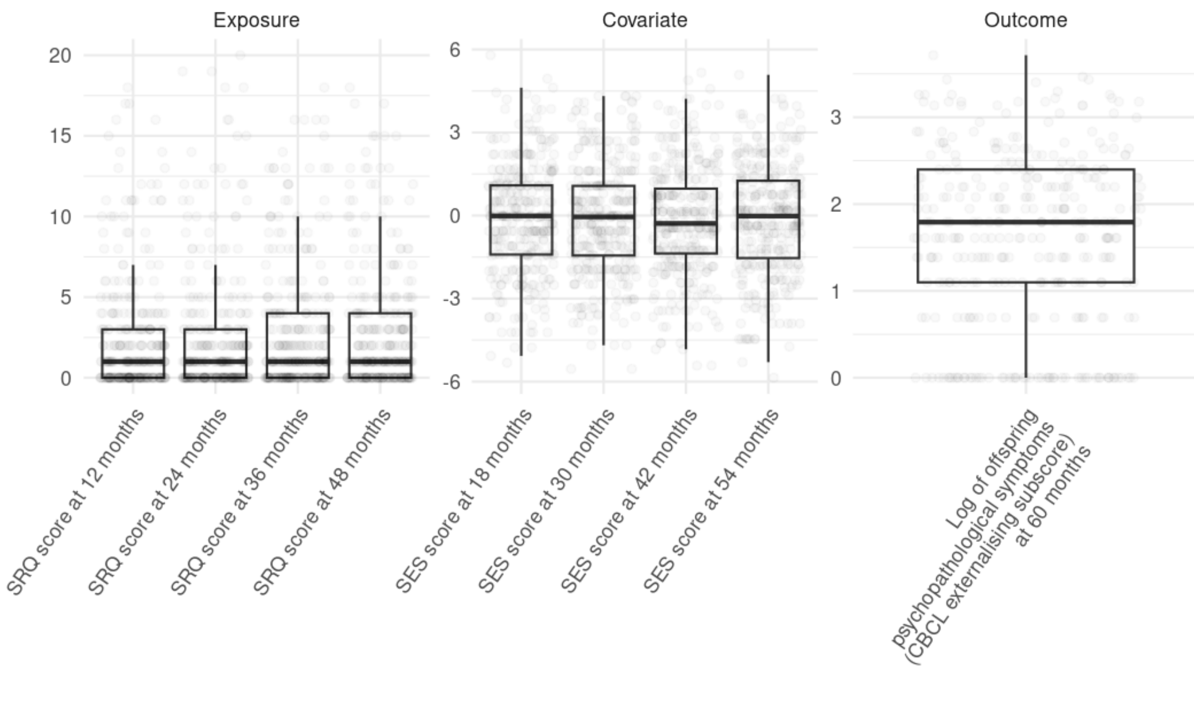

B)

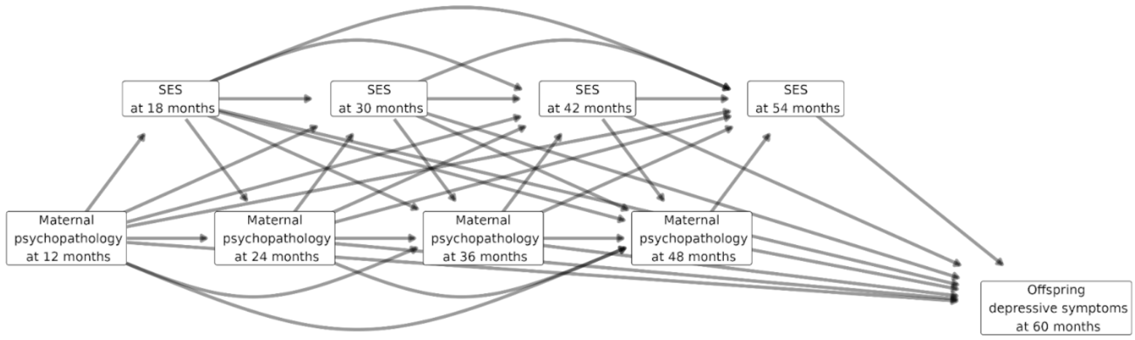

**Figure S2**. **A)** Distributions of exposure, covariate and outcome variables used in analyses. **B)** Causal DAG for empirical scenario with maternal psychological distress (SRQ score) as exposure, socioeconomic status (SES) as covariate, and log of offspring psychopathological symptoms (CBCL externalising subscore) as outcome.

| Variable | n | Mean | Median | Sd | Min | Max |
|---|---|---|---|---|---|---|
| SRQ score at 12 months | 336 | 2.40 | 1.00 | 3.60 | 0.00 | 18.00 |
| SRQ score at 24 months | 336 | 2.32 | 1.00 | 3.86 | 0.00 | 20.00 |
| SRQ score at 36 months | 336 | 2.65 | 1.00 | 3.71 | 0.00 | 18.00 |
| SRQ score at 48 months | 336 | 2.65 | 1.00 | 3.70 | 0.00 | 18.00 |
| SES score at 18 months | 336 | -0.10 | -0.01 | 1.97 | -5.32 | 5.79 |
| SES score at 30 months | 336 | -0.16 | -0.05 | 1.98 | -5.54 | 4.32 |
| SES score at 42 months | 336 | -0.19 | -0.29 | 1.95 | -5.42 | 5.16 |
| SES score at 54 months | 336 | -0.17 | -0.02 | 2.00 | -5.86 | 5.08 |
| Log of offspring psychopathological symptoms | 336 | 1.64 | 1.79 | 0.97 | 0.00 | 3.71 |

| |
|---|
| (CBCL externalising subscore) at 60 months |

**Table S1**. Summary statistics of exposure, covariate and outcome variables used in analyses.

#### Marginal structural model application

Marginal structural models, as described in Gilsanz et al. 2022[16], offer a complementary causal approach to identifying temporal effects of a repeatedly measured exposure over time in the presence of time-varying covariates, that may act as a mediator or a confounder. In such models, Inverse Probability of Treatment (or exposure) Weights (IPTW) allow the construction of a pseudo-population in which confounders are distributed identically across both exposed and unexposed individuals. The exposure, maternal psychopathology, measured using the SRQ score, was dichotomised with individuals scoring 6 and above considered exposed (1) and those below 6 considered unexposed (0). In **Model S1** $X_1$, $X_2$, $X_3$ and $X_4$ represent the exposure measured at 12, 24, 36 and 48 months of age respectively. The log of offspring depression, measured at 60 months of age using the CBCL score, was the outcome. The time-varying covariate, socio economic status, was adjusted for using stabilised IPTW. The denominator was calculated using logistic regression of a binary exposure on all variables measured earlier in time, to estimate the probability of an individual being exposed given their exposure and covariate history, respectively for each time point of exposure. The numerator was calculated using logistic regression of an exposure on the previous exposure, to estimate the probability of being exposed due to exposure in the previous exposure. The weight for each individual was calculated as the product of the numerators over the product of denominators. Confidence intervals were estimated through bootstrapping. The exponent of the coefficients $\theta_i$ is taken to obtain multiplicative factors for the effect of being exposed at time *i* on offspring depression given an individual's exposure and covariate history. The estimands obtained using this method differ in definition to those from DME SLCMA, representing the causal effect of intervening on every individual's exposure at a specific time versus not intervening, whilst holding all other exposures fixed. The **Model S1** assumes no interactions between exposures in their effects on the outcome. These are hypothetical interventions where it is assumed possible to intervene at a time point and set the maternal psychopathology state as exposed (1) or unexposed (0) without altering the status at any of the other time points. A more plausible intervention could be therapy or treatment; however, such treatment would not be randomly assigned in a birth cohort study, nor the therapy / treatments being identical for each recipient. Additionally, some individuals may not be able to receive therapy or treatment, such as for financial reasons or lack of availability of such a service.

$$E[Y(x_1, x_2, x_3, x_4)] = \theta_0 + \theta_1 x_1 + \theta_2 x_2 + \theta_3 x_3 + \theta_4 x_4 \qquad (S1)$$

| Maternal psychopathology exposure (binary) at: | Effect estimate | 95% lower | 95% upper |
|---|---|---|---|

| | | | |
|---|---|---|---|
| 12 months of age | 1.147 | 0.7771 | 1.6035 |
| 24 months of age | 0.921 | 0.5951 | 1.4724 |
| 36 months of age | 1.118 | 0.7472 | 1.6027 |
| 48 months of age | 1.587 | 1.1037 | 2.2559 |

**Table S2.** Effect estimates obtained from marginal structural model S1. Bootstrapped confidence intervals.

Counter factual definitions for life course hypothesis comparisons:

$$E[Y(1, x_2, x_3, x_4) - Y(0, x_2, x_3, x_4)] = \theta_1$$
$$E[Y(x_1, 1, x_3, x_4) - Y(x_1, 0, x_3, x_4)] = \theta_2$$
$$E[Y(x_1, x_2, 1, x_4) - Y(x_1, x_2, 0, x_4)] = \theta_3$$
$$E[Y(x_1, x_2, x_3, 1) - Y(x_1, x_2, x_3, 0)] = \theta_4$$

To compare the X4 critical period hypothesis with an accumulation hypothesis we used the marginal structural **Model S2**. This model tests if exposure accumulated over $X_1$ - $X_3$ affects the outcome, rather than just exposure at $X_4$. Here $\bar{x}$ denotes the exposure history shorthand for $(x_1, x_2, x_3, x_4)$ . We fit Model S2 by IPTW, assuming all causal assumptions hold and for binary exposures. This marginal structural model also assumes that the effect of the fourth exposure $x_4$ does not depend on the sum of the previous three exposures. For scenarios where this would be a possibility an interaction term may be included in the marginal structural model.

$$E[Y(\bar{x})] = \lambda_0 + \lambda_1 \sum_{i=1}^{3} x_i + \lambda_2 x_4 \qquad (S2)$$

$$E[Y(1,1,1, x_4)] - E(Y(0,0,0, x_4)] = 3\lambda_1$$

$$E[Y(x_i: \sum_{i=1}^{3} x_i = k\,, 1)] - E[Y(Y(x_i: \sum_{i=1}^{3} x_i = k\,, 0)] = \lambda_2$$

The coefficient $3\lambda_1$cam be interpreted as the average cumulative effect of setting the first three exposures to one, versus setting the first three exposures to zero, while holding the fourth exposure fixed. The coefficient $\lambda_2$ can be interpreted as the average effect of setting the fourth exposure to one versus setting the fourth exposure to zero, while holding the sum of the previous three exposures fixed. As shown in **Table S3**, exposure accumulated over $X_1$ - $X_3$ does not have a significant effect on the outcome, suggesting the $X_4$ critical period hypothesis better explains the outcome than the accumulation hypothesis.

| | Effect estimate | 95% lower | 95% higher |
|---|---|---|---|
| $\lambda_1$ | 1.051 | 0.914 | 1.186 |
| $\lambda_2$ | 1.617 | 1.157 | 2.424 |

**Table S3.** Effect estimates obtained from marginal structural model S2. Bootstrapped confidence intervals.

These marginal structural models provide complementary evidence on the relative magnitudes of total effects under different intervention timings, which are consistent with the findings of DME SLCMA, and provide fully specified causal estimands.

#### Post selection inference

Following the critical period at 48 months hypothesis selection by DME SLCMA, post selection inference was conducted using the max-|t| test[9,10], implemented using the SLCMA R package, accessible at *github.com/thedunnlab/slcma*[11–13].

| Method | Effect Estimate | Confidence interval lower (95%) | Confidence interval upper (95%) | Adj. *P*-value |
|---|---|---|---|---|
| Bonferroni | 1.039 | 1.010 | 1.069 | 0.039 |
| Max-\|t\| | 1.039 | 1.003 | 1.076 | 0.028 |

**Table S4**. Post selection inference results across methods.

#### Deterministic simulation study

| Factor | Levels | Description |
|---|---|---|
| Correlation structure | 1. Observed<br>2. Increasing<br>3. Decreasing<br>4. Exposure to covariate greater | 1. Correlation structure in an empirical DCHS data set<br>2. Correlation between variables increases over time<br>3. Correlation between variables decreases over time |

| | | |
|---|---|---|
| | 5. Covariate to exposure greater | 4. Exposure to following covariate greater than covariate to following exposure<br>5. Covariate to following exposure greater than exposure to following covariate |
| Exposure direct effect | 1. X1<br>2. X2<br>3. X3<br>4. A<br>5. None | 1. Exposure direct effect from X1 to Y<br>2. Exposure direct effect from X2 to Y<br>3. Exposure direct effect from X3 to Y<br>4. Exposure direct effect from all exposures to Y<br>5. No direct effects from exposures to Y |
| Covariate direct effect | 1. C1<br>2. C2<br>3. C3<br>4. All<br>5. None | 1. Covariate direct effect from C1 to Y<br>2. Covariate direct effect from C2 to Y<br>3. Covariate direct effect from C3 to Y<br>4. Covariate direct effect from all covariates to y<br>5. No direct effects from exposures to Y |
| Covariate direct effect size | 1. 0.05<br>2. 0.1<br>3. 0.2<br>4. 0.5 | 1. Small covariate direct effects<br>2. Medium covariate direct effects<br>3. Large covariate direct effects<br>4. Extreme covariate direct effects |
| Correlation strength | 1. 0<br>2. 1<br>3. 2<br>4. 25 | A nonlinear parameter that controls the correlation levels between exposures and covariates without affecting the correlation levels within covariates and exposures.<br>1. No exposure-covariate correlation<br>2. Default correlation structure exposure-covariate correlation<br>3. ~2x default correlation structure exposure-covariate correlation<br>4. Extreme exposure-covariate correlation |

**Table S5**. Deterministic simulation study factor names and description, and the respective levels of each.

A)

B)

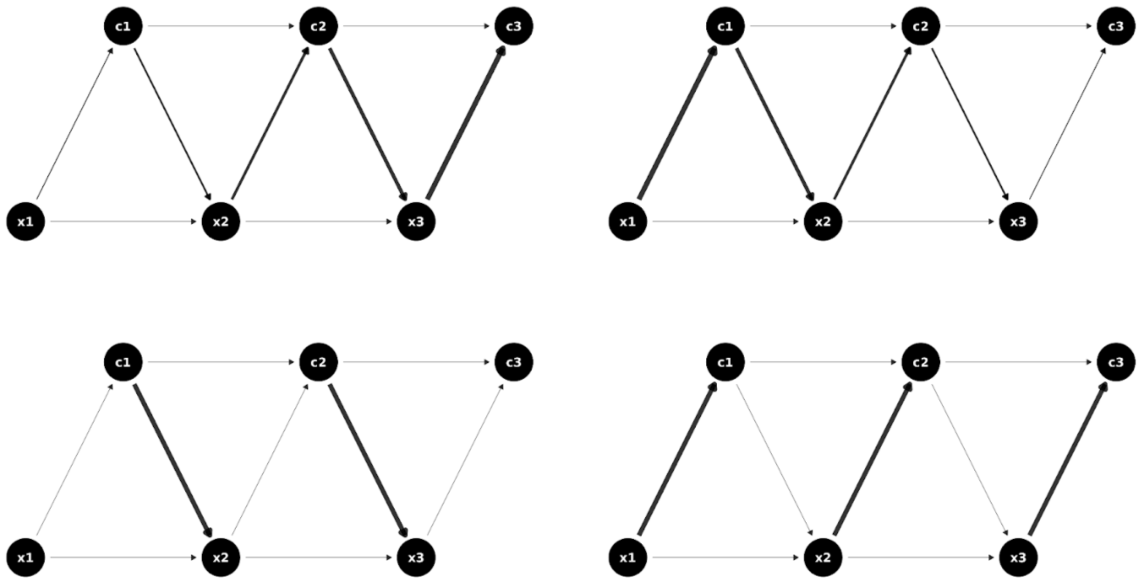

**Figure S3**. **A)** Increasing and decreasing correlation structures. **B)** Covariate to exposure greater and exposure to covariate greater correlation structures.

Three time points of exposure and covariate data for 500 individuals were simulated from a standard normal distribution, for each scenario, with a covariance structure for each correlation structure imposed using Cholesky decomposition. The simulated exposures and covariates were used to deterministically calculate the outcome variable using the respective parameter factors for each scenario, from the equation

$$y = \beta_1 x_1 + \beta_2 x_2 + \beta_3 x_3 + \beta_4 c_1 + \beta_5 c_2 + \beta_6 c_3,$$

where $\beta_i$ is the direct effect magnitude for variable $i$ on the outcome.

#### Deterministic simulation study results

A)

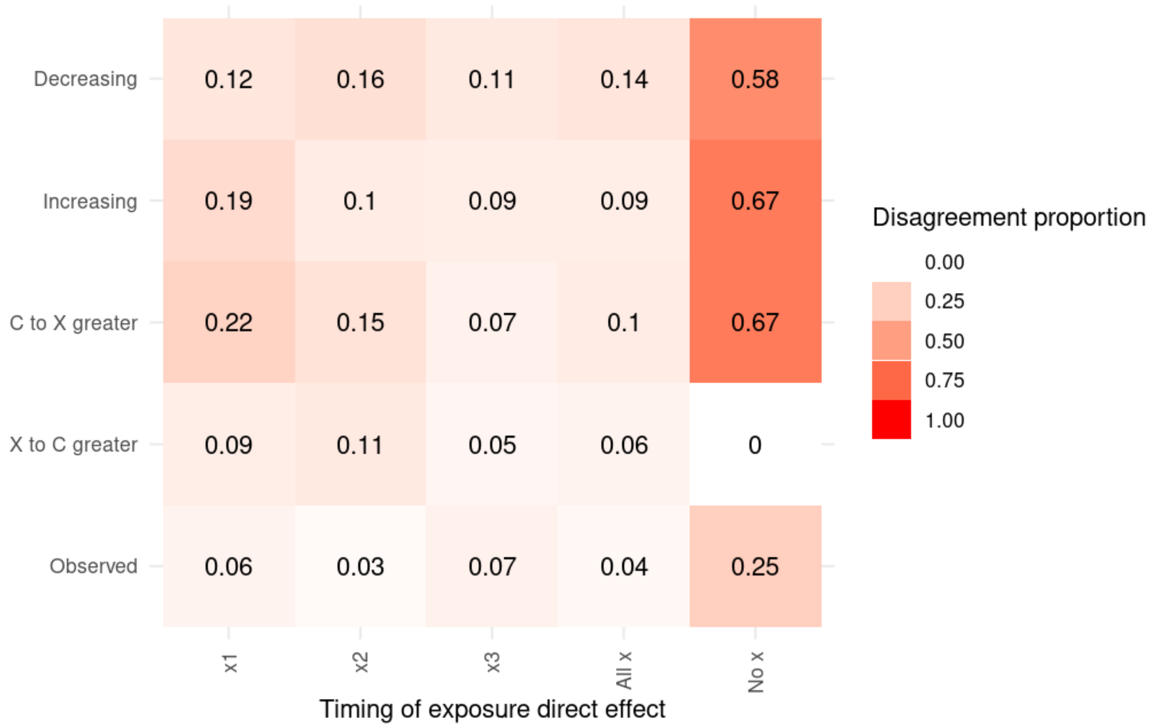
Decreasing
Increasing
C to X greater
X to C greater
Observed
0.12 0.16 0.11 0.14 0.58
0.19 0.1 0.09 0.09 0.67
0.22 0.15 0.07 0.1 0.67
0.09 0.11 0.05 0.06 0
0.06 0.03 0.07 0.04 0.25
x1
x2
x3
All x
No x
Timing of exposure direct effect
Disagreement proportion
0.00
0.25
0.50
0.75
1.00

B)

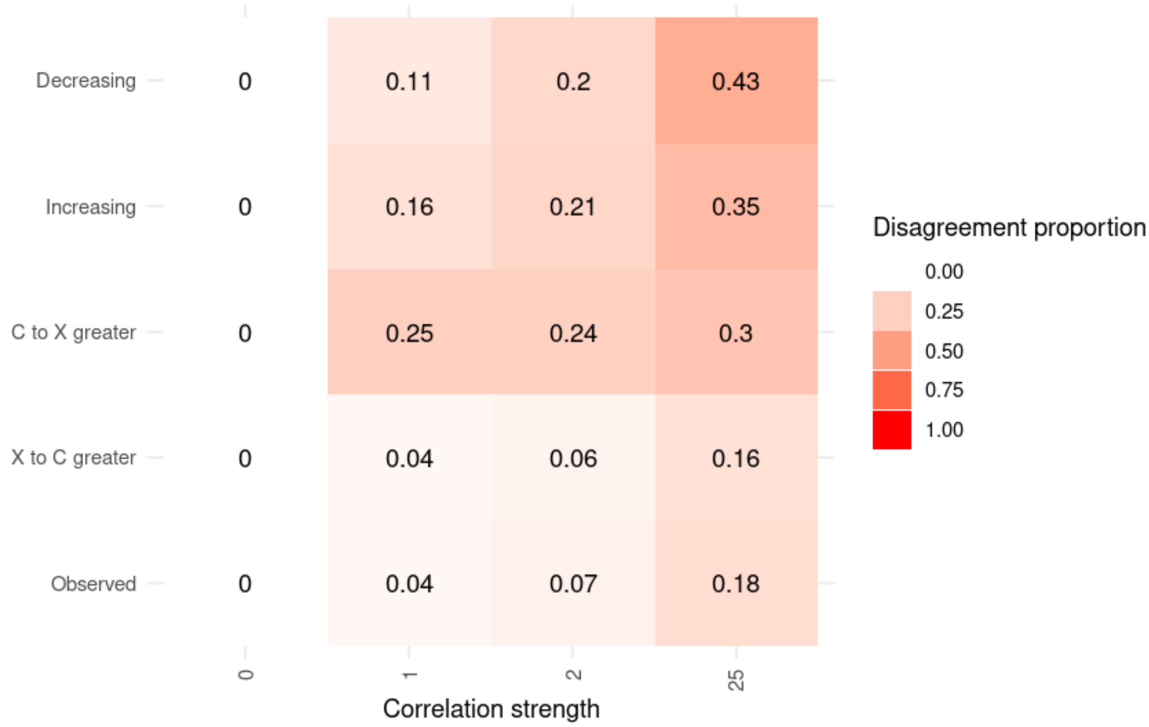
Decreasing
Increasing
C to X greater
X to C greater
Observed
0 0.11 0.2 0.43
0 0.16 0.21 0.35
0 0.25 0.24 0.3
0 0.04 0.06 0.16
0 0.04 0.07 0.18
0
1
2
25
Correlation strength
Disagreement proportion
0.00
0.25
0.50
0.75
1.00

C)

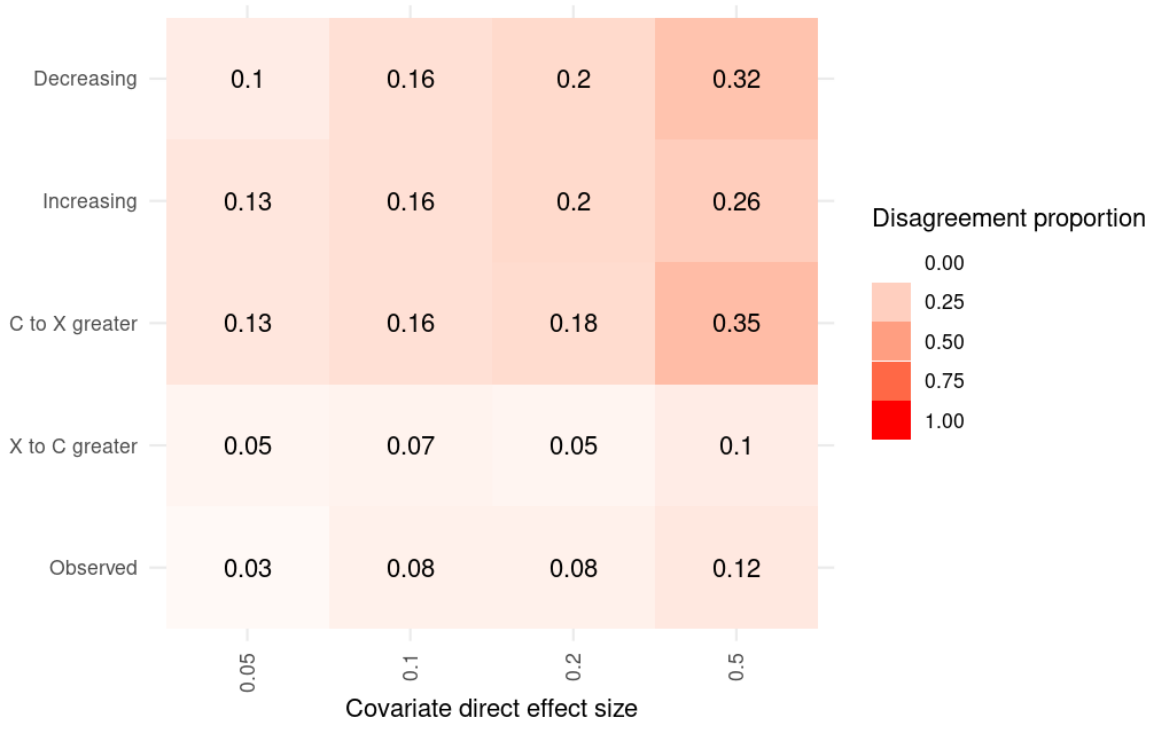

D)

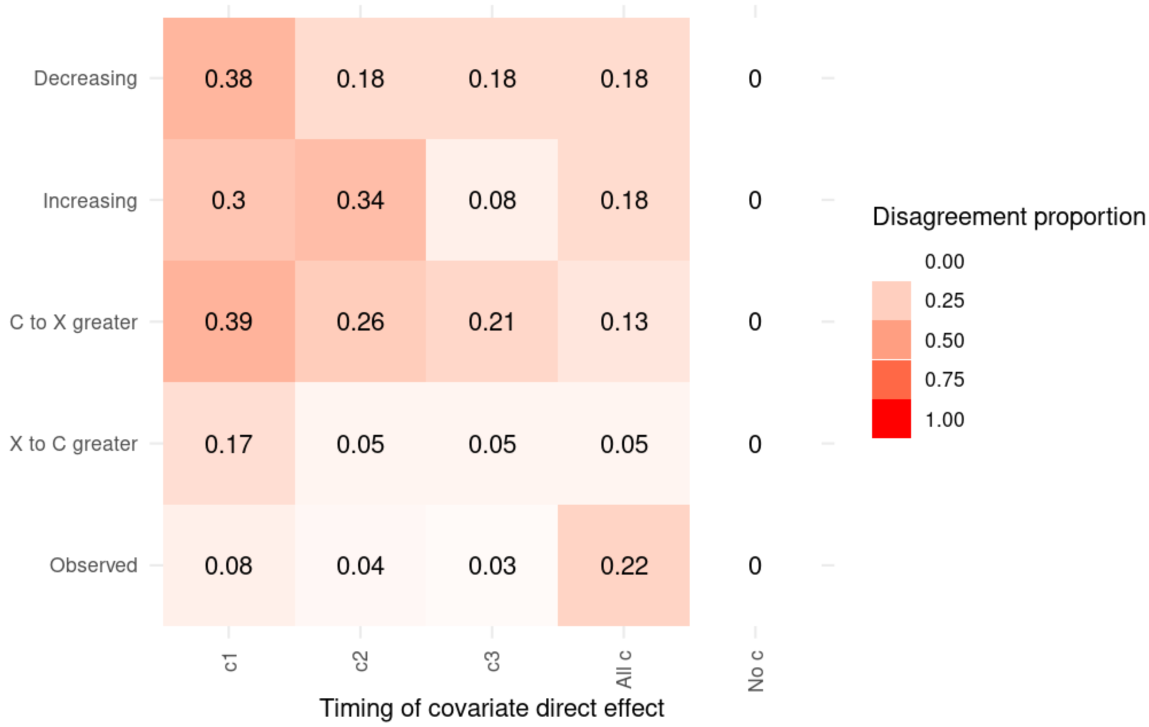

**Figure S4**: Second level factor plots from the deterministic simulation study showing how each factor interacts with different correlation structure. Each tile shows the proportion of scenarios where regular SLCMA and DME SLCMA disagree on the hypothesis selected across all scenarios with the respective factor levels. **A)** Correlation structures vs Exposure direct effect timing. **B)** Correlation structures vs Exposure-covariate correlation strength. **C)**

Correlation structures vs Covariate direct effect size. **D)** Correlation structures vs Covariate direct effect timing.

**Figure S4** highlights how the presence of confounding, when regular SLCMA disagrees in hypothesis selection with DME SLCMA, depends on a number of factors, and their interaction. **Figure S4A** shows that in general confounding is much more likely to be present when there are no direct exposure effects on the outcome, with the exception of the X to C greater correlation structure where there was no disagreement between methods when there were no direct exposure effects present. The X to C greater correlation structure increases the relative strength of mediated exposure effects to the confounding covariate effect pathways.

**Figure S4B** indicates that confounding can be present at lower levels of exposure-covariate correlation depending on the correlation structure of the exposures and covariates. At the observed levels of exposure-covariate correlations 25% of the C to X greater correlation structures scenarios displayed disagreement between the methods, considerably larger than the other correlation structures. The C to X greater correlation structure increases the relative strength of the confounding covariate effect pathways to the mediated exposure effects.

**Figure S4C** shows a similar result to **Figure S4B**, the Observed and X to C greater correlation structures have a lesser increase in disagreement between methods, in comparison to the other correlation structures, when the covariate direct effect size, which has an effect on the confounding pathways, is increased.

**Figure S4D** shows how the confounding effect of the timing of covariate effects on the outcome is heavily dependent on the exposure-covariate correlation structure, with the correlation structures being most susceptible to confounding from direct covariate effects at different time-points. The Observed correlation structure is most susceptible to confounding when there are covariate direct effects present at all time-points, the Increasing correlation structure when there is an effect from the middle covariate time-point and the other correlation structures when there is a covariate direct effect from the first time-point. In general, the covariate direct effects earlier in time cause more confounding than that those later as more backdoor paths are present from earlier covariate direct effects.

#### Stochastic Simulation study

| Factor | Levels | Description |
|---|---|---|
| Correlation structure | 1. Observed (Baseline)<br>2. Increasing<br>3. Decreasing | 1. Correlation structure in an empirical DCHS data set<br>2. Correlation between variables increases over time |

| | | |
|---|---|---|
| | 4. Exposure to covariate greater<br>5. Covariate to exposure greater | 3. Correlation between variables decreases over time<br>4. Exposure to following covariate greater than covariate to following exposure<br>5. Covariate to following exposure greater than exposure to following covariate |
| Exposure direct effect | 1. X1<br>2. X2 (Baseline)<br>3. X3<br>4. A<br>5. None | 1. Exposure direct effect from X1 to Y<br>2. Exposure direct effect from X2 to Y<br>3. Exposure direct effect from X3 to Y<br>4. Exposure direct effect from all exposures to Y<br>5. No direct effects from exposures to Y |
| Covariate direct effect | 1. C1<br>2. C2 (Baseline)<br>3. C3<br>4. All<br>5. None | 1. Covariate direct effect from C1 to Y<br>2. Covariate direct effect from C2 to Y<br>3. Covariate direct effect from C3 to Y<br>4. Covariate direct effect from all covariates to Y<br>5. No direct effects from exposures to Y |
| Covariate direct effect size | 1. 0.05<br>2. 0.1<br>3. 0.2 (Baseline)<br>4. 0.5 | 1. Small covariate direct effects<br>2. Medium covariate direct effects<br>3. Large covariate direct effects<br>4. Extreme covariate direct effects |
| Correlation strength | 1. 0<br>2. 1<br>3. 2 (Baseline)<br>4. 25 | A nonlinear parameter that controls the correlation levels between exposures and covariates without affecting the correlation levels within covariates and exposures.<br>1. No exposure-covariate correlation<br>2. Default correlation structure exposure-covariate correlation<br>3. ~2x default correlation structure exposure-covariate correlation<br>4. Extreme exposure-covariate correlation |

| | | |
|---|---|---|
| Sample size | 1. 100<br>2. 500 (Baseline)<br>3. 1000 | Number of individuals simulated in each iteration. |
| Noise | *1.* $R^2$ = 0.1 (Baseline)<br>*2.* $R^2$ = 0.01 | Proportion of outcome variance explained by exposure direct and mediated effects. |

**Table S6**. Stochastic semi-factorial simulation factor names and descriptions, and the respective levels of each.

The exposure and covariate data for a baseline of 500 individuals was simulated with 1000 iterations for each semi-factorial level using the same method as for the deterministic simulations. The outcome variable for each individual in each iteration was calculated using the equation

$$y_i = \beta_1 x_{1i} + \beta_2 x_{2i} + \beta_3 x_{3i} + \beta_4 c_{1i} + \beta_5 c_{2i} + \beta_6 c_{3i} + \epsilon_i,$$

where the residuals $\epsilon_i$ are normally distributed with the variance controlled such that the total causal effect of all exposure and covariates on the outcome has an $R^2$ of 0.1 at baseline. In addition to the factors used in the deterministic simulations, we introduce factors for sample size (*n = 100, 500, 1000*) and measurement error by varying $R^2$ (0.01, 0.1) to our semi-factorial simulation design. This gives a total of 21 scenarios to the stochastic simulation study: 1 baseline, 4 correlation structures, 4 exposure effect timings, 4 covariate effect timings, 3 covariate effect sizes, 2 levels of correlation strength (no exposure-covariate correlation not included), 2 sample sizes and 1 measurement error.

A)

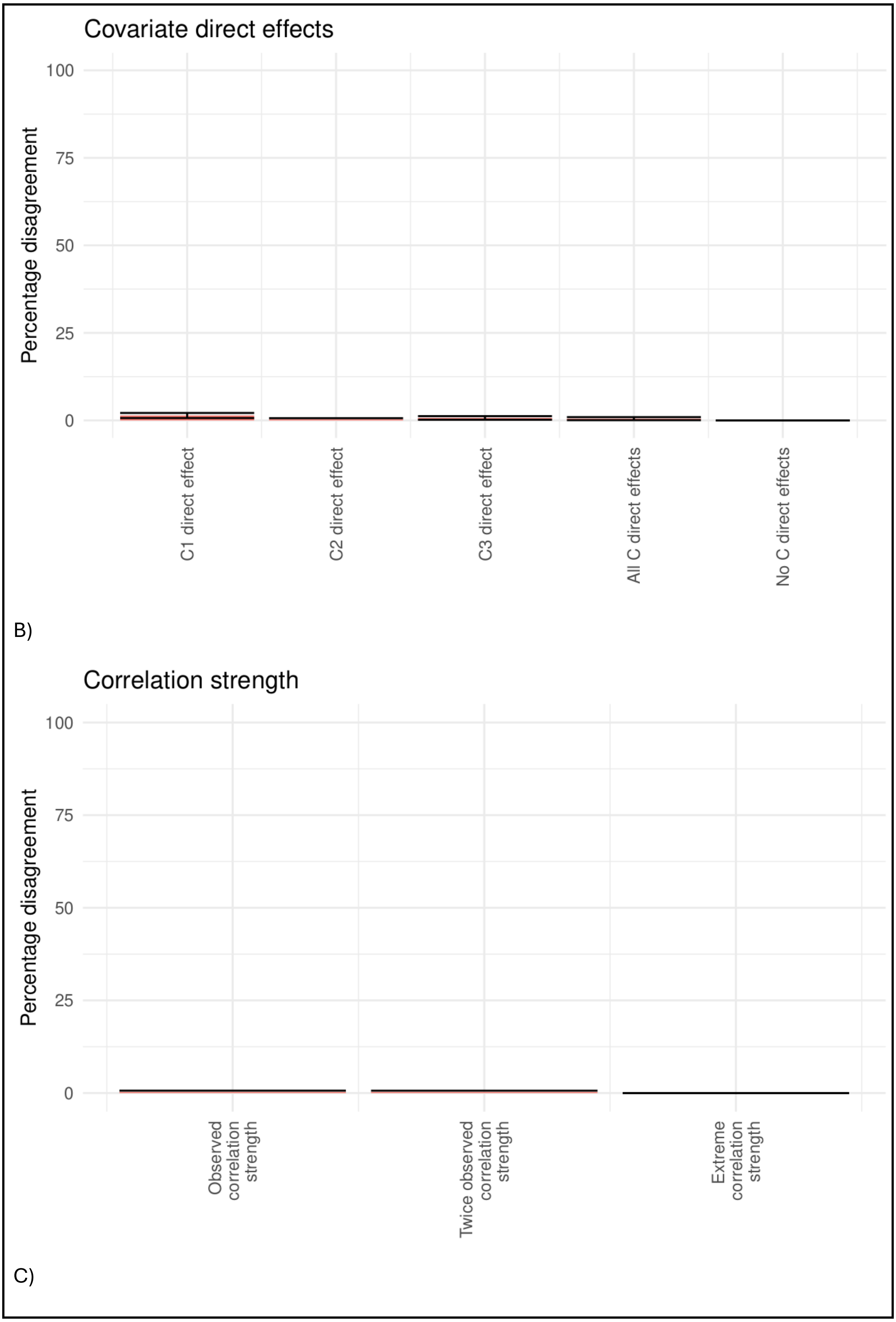
Covariate direct effects
Percentage disagreement
100
75
50
25
0
C1 direct effect
C2 direct effect
C3 direct effect
All C direct effects
No C direct effects
B)
Correlation strength
Percentage disagreement
100
75
50
25
0
Observed correlation strength
Twice observed correlation strength
Extreme correlation strength
C)

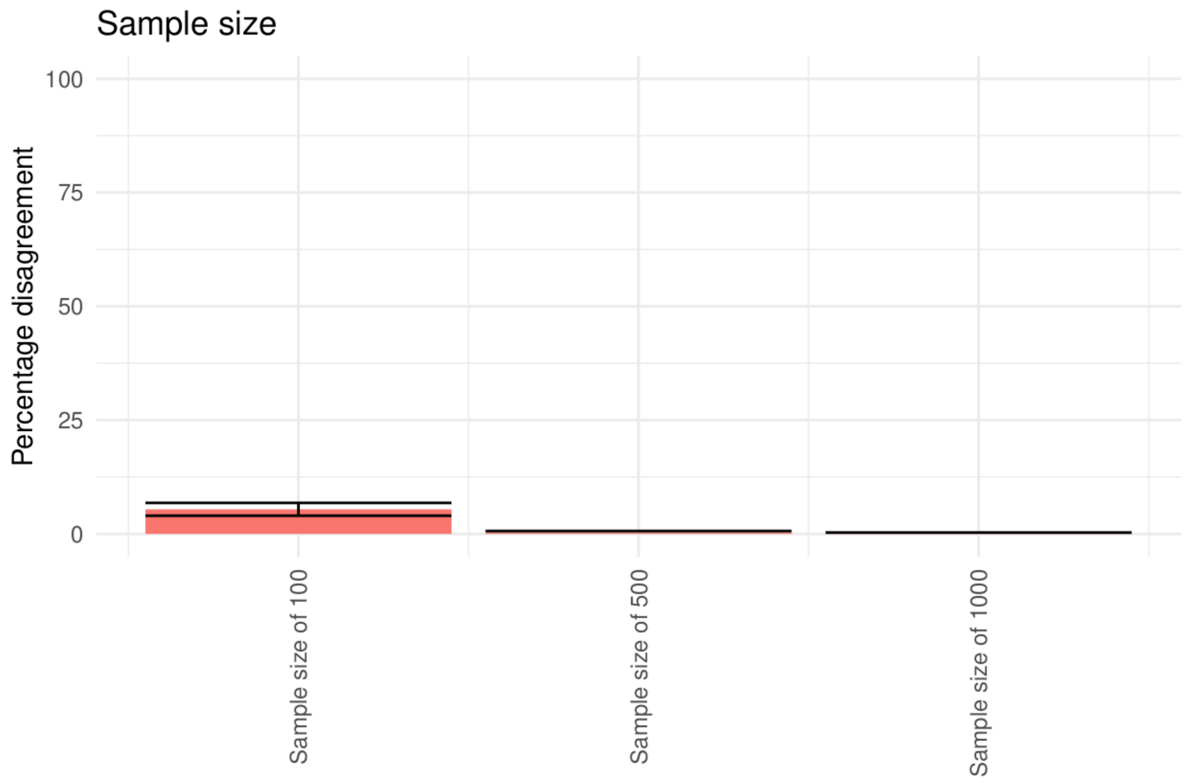

D)

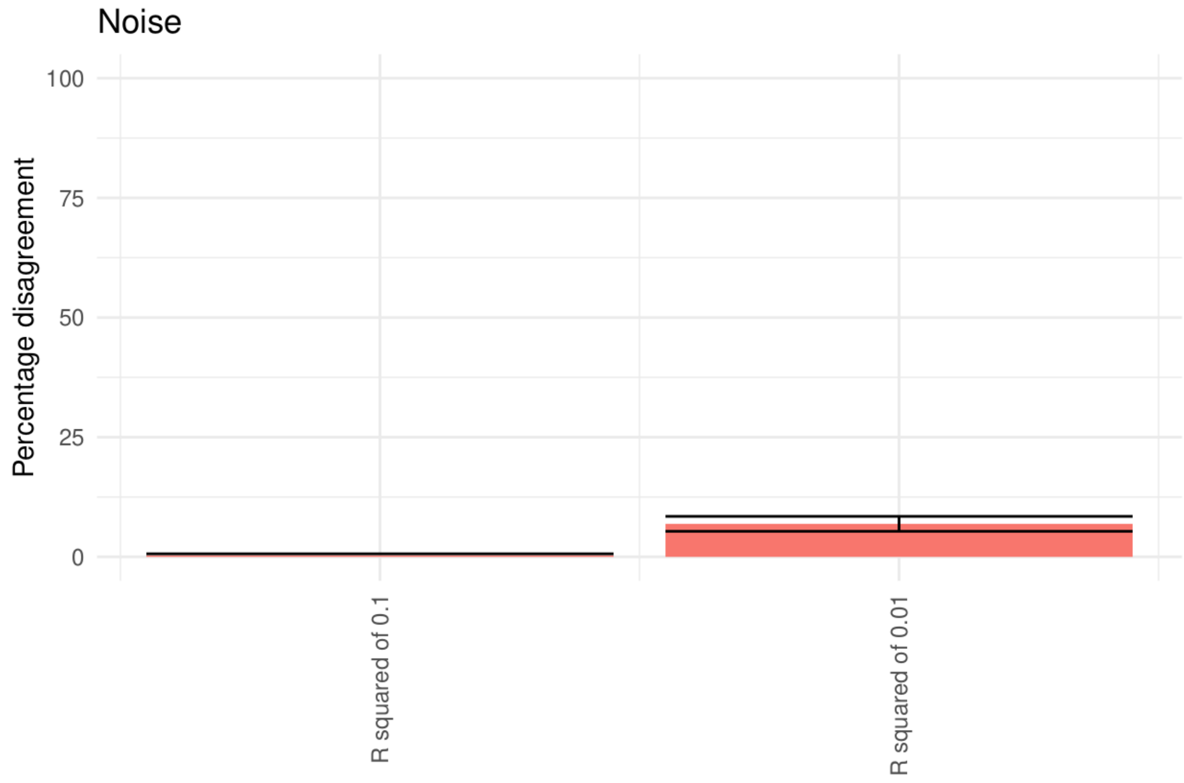

**Figure S5**. Stochastic simulation results. **A-D)** Percentage disagreement in 1000 iterations as factors are varied from baseline. **A)** Covariate direct effects timing **B)** Correlation strength **C)** Sample size **D)** Noise. Bootstrapped confidence intervals are displayed in plots **A-D.**

.

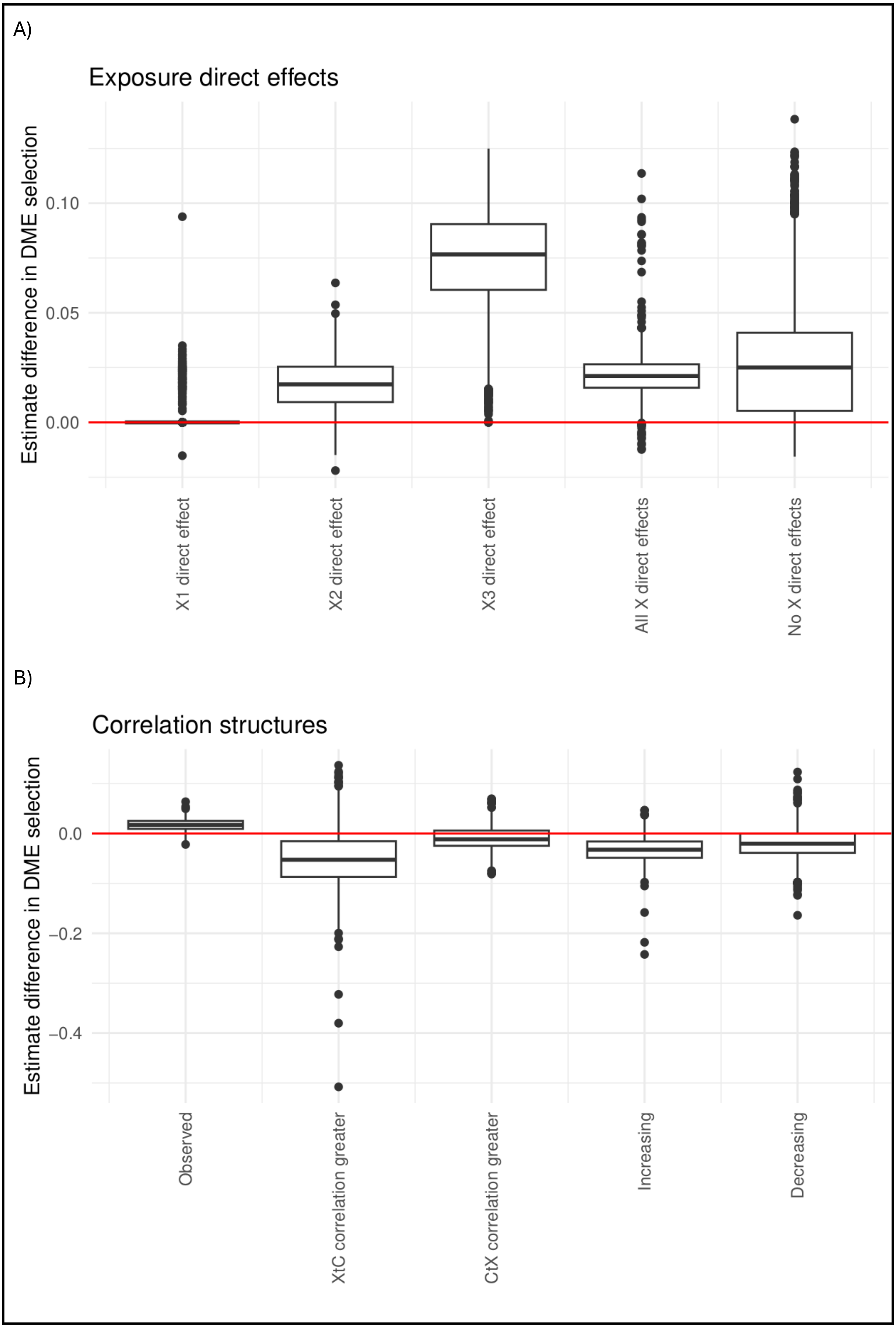
A)
Exposure direct effects
Estimate difference in DME selection
0.10
0.05
0.00
X1 direct effect
X2 direct effect
X3 direct effect
All X direct effects
No X direct effects
B)
Correlation structures
Estimate difference in DME selection
0.0
−0.2
−0.4
Observed
XtC correlation greater
CtX correlation greater
Increasing
Decreasing

C)

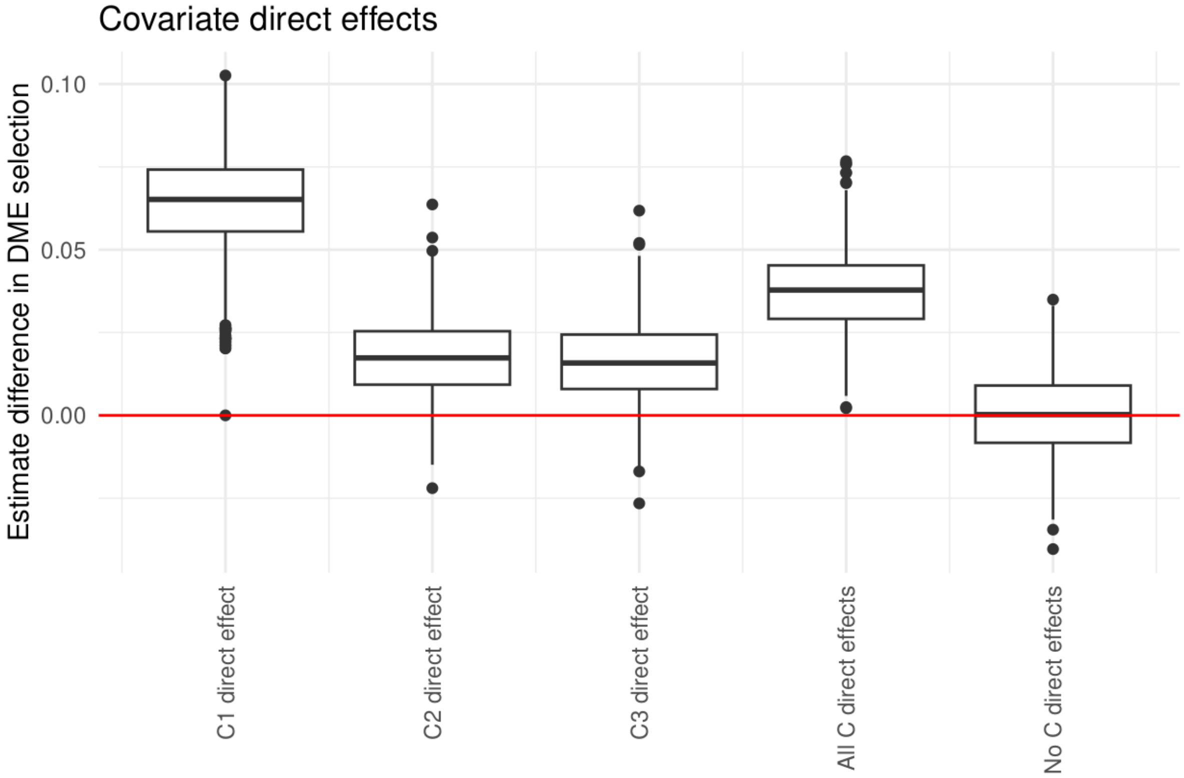
Covariate direct effects
Estimate difference in DME selection
0.10
0.05
0.00
C1 direct effect
C2 direct effect
C3 direct effect
All C direct effects
No C direct effects

D)

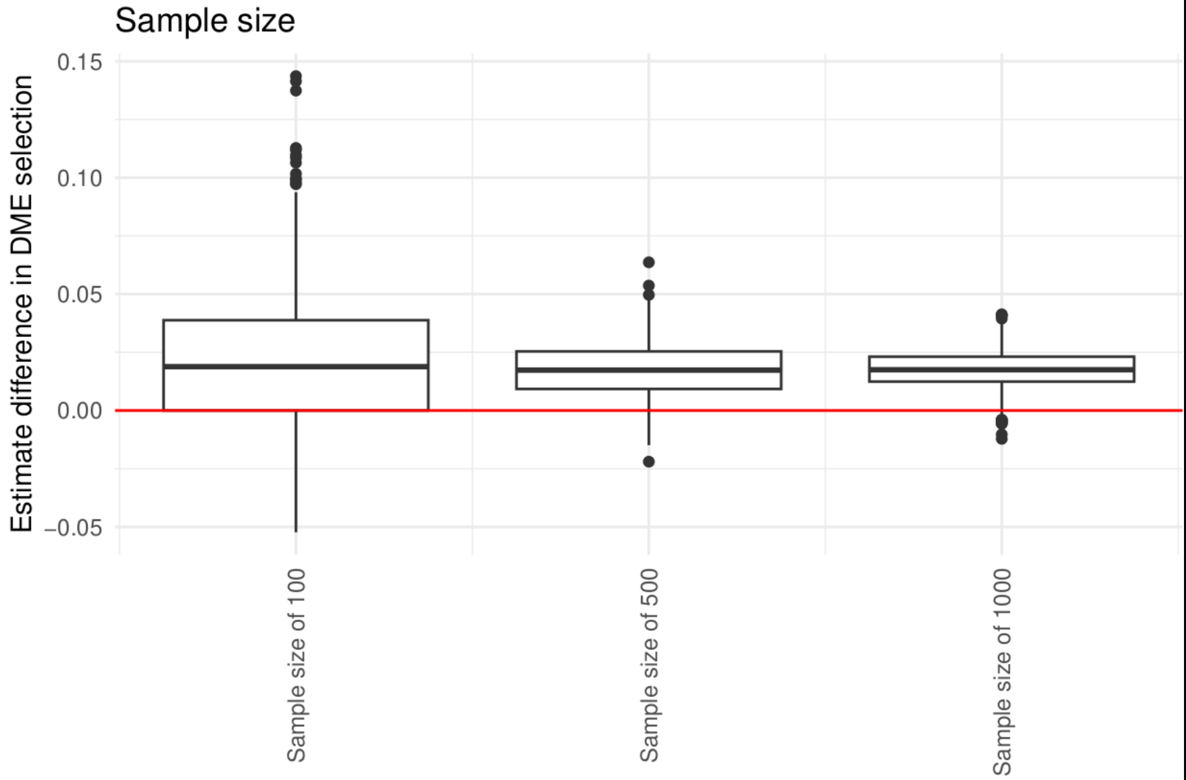
Sample size
Estimate difference in DME selection
0.15
0.10
0.05
0.00
−0.05
Sample size of 100
Sample size of 500
Sample size of 1000

E)

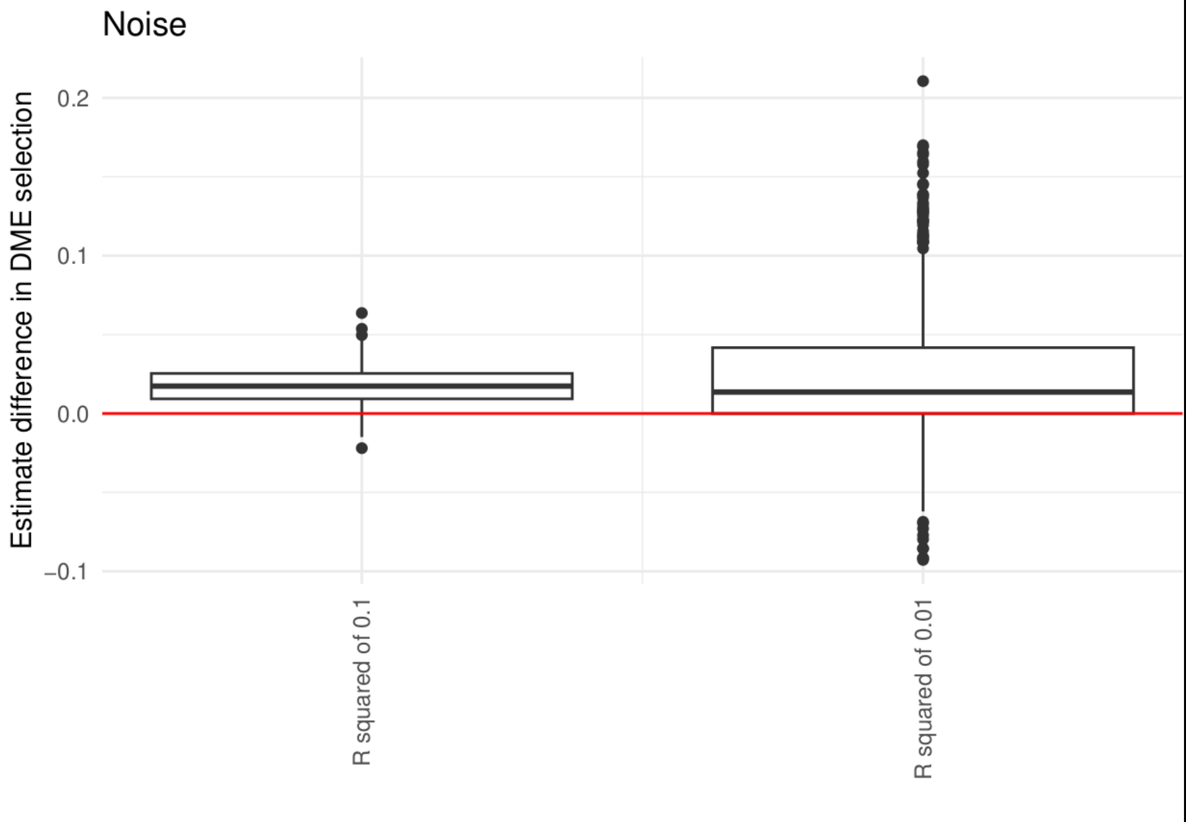

**Figure S6**. Stochastic simulation results. **A-E)** Differences in effect estimates between DME SLCMA and SLCMA for the DME SLCMA selected hypothesis.

#### Code availability

Code for the deterministic and stochastic simulation studies, empirical application and post selection inference is available on github: github.com/SEBeer1/DME_SLCMA
